# Trends in the prevalence of overweight and obesity in children and adolescents in France in the 21^st^ century: a systematic review and meta-analysis

**DOI:** 10.1101/2025.09.22.25336050

**Authors:** Marianne Jacques, Martin Chalumeau, Camille Le Gal, Benoit Salanave, Bruno Frandji, Marie-Aline Charles, Sandrine Lioret, EBGM IX study group, Barbara Heude, Pauline Scherdel

## Abstract

**Background:** We aimed to systematically assess temporal trends in paediatric overweight and obesity prevalence in France over the past 2 decades, given the lack of regular nationwide monitoring.

**Methods:** We systematically searched for both published and unpublished studies reporting the prevalence of overweight (including obesity) and obesity, defined by BMI and IOTF thresholds, from January 2000 to December 2024 in the paediatric population (aged 2-17 years) living in France. Risk of bias was assessed with a customised version of Hoy’s score for prevalence studies. We selected nationwide studies with a low risk of bias, stratified by sex, to estimate summary prevalence using meta-analyses and to assess temporal trends using meta-regressions adjusted for age, using inverse-variance–weighted, multilevel, linear, random-effects models. We conducted sensitivity analyses including all studies regardless of risk of bias.

**Findings:** Of 2,421 publications screened, 188 were included and 122 were unique studies; 18 studies (n=143,655 children) were nationwide with low risk of bias. No nationwide study at low risk of bias was available after 2017. Summary prevalence of overweight (including obesity) was 18.2% (95% CI 16.5-20.0) for girls and 16.0% (14.2-17.9) for boys. We found no statistically significant temporal trend for overweight and obesity from 2000 to 2017 for either sex. The sensitivity analyses revealed significant positive temporal trends for overweight and obesity, except for overweight in boys.

**Interpretation:** We found no significant increase in the prevalence of overweight and obesity among children and adolescents living in France from 2000 to 2017. However, including studies regardless of risk of bias led to different conclusions. Repeated, high-quality nationwide studies are urgently needed in France to reliably monitor trends in paediatric overweight and obesity prevalence and to assess the effectiveness of public health policy.

**Funding:** Doctoral School 393 Pierre Louis of Public Health (CLG).

## INTRODUCTION

Overweight and obesity in childhood and adolescence represent a major public health challenge throughout the world. Since 1990, obesity has more than doubled worldwide in adults and quadrupled in children and adolescents aged 5 to 19; high-income countries are not spared.^1–3^ Obesity increases the risk of the development of serious health complications such as chronic respiratory diseases, metabolic complications, cardiovascular diseases, musculoskeletal disorders, or cancers.^4^ In children and adolescents, it is also associated with adverse psychological disorders, including anxiety, depression, low self-esteem, and social isolation, often linked to weight-related stigma.^5, 6^ Since the early 2000s, the World Health Organisation (WHO) has launched several plans to tackle obesity, the last one being the 2022 Acceleration Plan to Stop Obesity.^7, 8^ These plans provide structured methodologies to implement preventive policies tailored to national contexts, along with analytic tools to evaluate their effectiveness. Such evaluation requires a rigorous monitoring system based on a regularly repeated and unbiased national assessment of the prevalence of paediatric overweight and obesity. This evaluation can be challenging owing to the many potential sources of bias in studies of weight and height, particularly selection bias and reliance on self-reported data.^9^ Reaching children and adolescents from socially disadvantaged families, who are disproportionally affected by overweight,^10, 11^ and engaging them in surveys is more difficult than studying children and adolescents from advantaged families, which contributes to selection bias and likely leads to an under-estimation of obesity prevalence.

In many high-income countries, such as England^12^ and the United States,^13^ regular surveys on the prevalence of overweight and obesity are available because of nationwide studies with low risk of bias. These surveys allow for monitoring the effectiveness of public health policies. Among high-income countries, France has a relatively unique situation regarding the epidemic of paediatric overweight and obesity. First, using the definition of the International Obesity Task Force (IOTF),^14, 15^ the prevalence of paediatric overweight (including obesity) was estimated at 15% to 20% in the early 2000s, which is lower than estimates in some other high-income countries (20% to 25%).^16, 17^ Second, although several national strategies to reduce paediatric overweight and obesity, such as the French National Nutrition and Health Program,^18^ have been in place since 2001, there is still no regular and rigorous monitoring of the prevalence of overweight and obesity.

We aimed to assess the temporal trends in the prevalence of paediatric overweight and obesity in France by using a systematic review and meta-analysis, including a careful analysis of the impact of risk of bias in the included studies.

## METHODS

### General methodology

This systematic review was conducted^19^ and reported^20^ according to the preferred reporting items for systematic reviews and meta-analyses statement (**Supplemental Table 1**). Two co-authors (MJ and CLG) independently performed the database searches, screened articles for inclusion, extracted and synthesised data, and assessed risk of bias; disagreements were resolved by consensus or by consulting other review authors (PS, BH).

### Search strategy and selection criteria

We searched for study reports published from January 1, 2000 to December 31, 2024 in MEDLINE via PubMed and Web of Science electronic databases. The search strategy combined keywords pertaining to obesity, overweight and childhood with a geographic restriction to France (**Supplemental Table 2**). No language restrictions were applied. We also searched Google Scholar for studies citing included studies and examined the first 20 (arbitrary) related articles of included studies in PubMed. Other reports on overweight and obesity in French children and adolescents were identified by a search of international, regional and national health agency websites (see list in **Supplemental Table 3**).

We included cross-sectional, longitudinal, or interventional studies reporting the prevalence of overweight (including obesity) and obesity, defined by body mass index (BMI) and IOTF thresholds, in the general population of children and adolescents aged 2 to 17 years in France. Several BMI growth charts exist around the world and the two most commonly used at national and international levels are those from the WHO^21^ and IOTF.^14, 15^ The latter growth chart has the advantage of offering continuity with the definition of adult overweight and obesity at age 18 and has been recommended in France since its publication by the French health authority in the 2000s, which is why we chose it.^18^ We excluded studies that targeted a specific subgroup of the population concerning its socioeconomic position or chronic conditions (e.g., neurological disorders).

### Data extraction and synthesis

We included eligible articles by reading their titles and abstracts, then their full text, if needed. We extracted the following data from included studies: characteristics of the study (design, location based on the Nomenclature of Territorial Units for Statistics [NUTS] of the European Union, participation rate, and methods used for anthropometric measurement), the population (calendar year interval, age interval, sex, and sample size), and the prevalence of overweight (including obesity) and obesity. For intervention studies, we extracted prevalence at baseline. When data were missing or unclear, the authors of the articles were contacted at least three times.

### Risk-of-bias assessment

The evaluation of the risk of bias of included studies was based on the checklist proposed by Hoy *et al* and Munn *et al* for prevalence studies,^22–24^ customised to our research question. Studies were considered at low risk of bias if all of the following criteria were fulfilled: (i) the sampling design allowed for providing results representative of the general population of the age group and area studied, (ii) the children and adolescents were recruited exhaustively or randomly from the source population, (iii) inappropriate exclusions were avoided (e.g., exclusion based on child or family characteristics), (iv) initial refusal rate to participate was low (arbitrarily defined as below the first quartile of the overall refusal rates among included studies) or appropriately addressed (by weighting), and (v) anthropometric measurements were accurate (e.g., standardised measurement, with individuals wearing light clothes and no shoes, using a scale and height gauge with low measurement inaccuracy) (**Supplemental Table 4**). Studies were considered at unclear risk of bias if ≥1 of the above criteria were judged unclear and the other criteria were judged at low risk of bias. Studies were considered at high risk of bias if they did not meet ≥1 of the above criteria.

### Data analysis

The statistical unit used was the study by calendar year interval, age interval, and sex. We performed the main analyses with only nationwide studies (recruitment over mainland France, including or not overseas departments and regions) considered at low risk of bias. The main outcomes were the prevalence of overweight (including obesity) and obesity. The distribution of prevalence was examined and, if necessary, transformed by using the Freeman-Tukey double arcsine.^25^ We calculated the midrange of the calendar year interval and the age interval for each study. The calendar year midrange was centred on the minimum calendar year (baseline: 2000) and was examined as a continuous variable. The age midrange was examined as a categorical explanatory variable divided into three classes: 2 to <6, 6 to <11, and 11 to <18 years.

First, we conducted meta-analyses using inverse-variance–weighted, multilevel, linear, random-effects models with and without stratification on sex to estimate the summary prevalence and their 95% confidence interval (95% CIs) from 2000 to 2024, globally and by age class. We examined the proportion of variability attributable to between-study heterogeneity using Cochran’s Q test (with p <0·05 indicating the presence of heterogeneity) and Higgins I^2^ statistic (with 0% indicating no heterogeneity and 100% large heterogeneity).^26, 27^ To obtain a summary prevalence over the period, we used a fixed-effects model when heterogeneity was low or a random-effects model otherwise.

Second, we conducted meta-regressions using inverse-variance–weighted, multilevel, linear, random-effects models to assess temporal trends in prevalence over time with adjustment for age class and stratification by sex. We included an interaction term between calendar year midrange, age class, and sex if significant (p <0.20) to account for potential age- and sex-specific BMI trajectories during childhood.^28^ We plotted the modelled prevalence by calendar year midrange and age class.

We performed two sensitivity analyses. First, we estimated the summary prevalence, globally and by age class, and assessed the temporal trends in prevalence over time using data from all studies including those at unclear and at high risk of bias. Second, we transformed the prevalence, if necessary, using the logit transformation. For all analyses, we used R V.4.3.3 (R Foundation for Statistical Computing, Vienna, Austria).

## RESULTS

### Results of the search and identification of studies

We identified 2,421 potentially eligible publications. After screening titles and abstracts, we assessed 198 full publications for eligibility and included 87. We included 101 additional reports retrieved from health agency websites (**Figure 1**). Finally, we included reports of 122 unique studies, of which 29% were nationwide (n=35, including 24 conducted at the nationwide level only in mainland France), 37% (n=45) were at the NUTS 2 level, 18% (n=22) the NUTS 3 level, and 16% (n=20) the local administrative level (**Supplemental Figure 1**). Several studies did not report results stratified by sex (n=3) or did not report results for overweight (including obesity) (n=22) or obesity (n=24).

**Figure 1.**
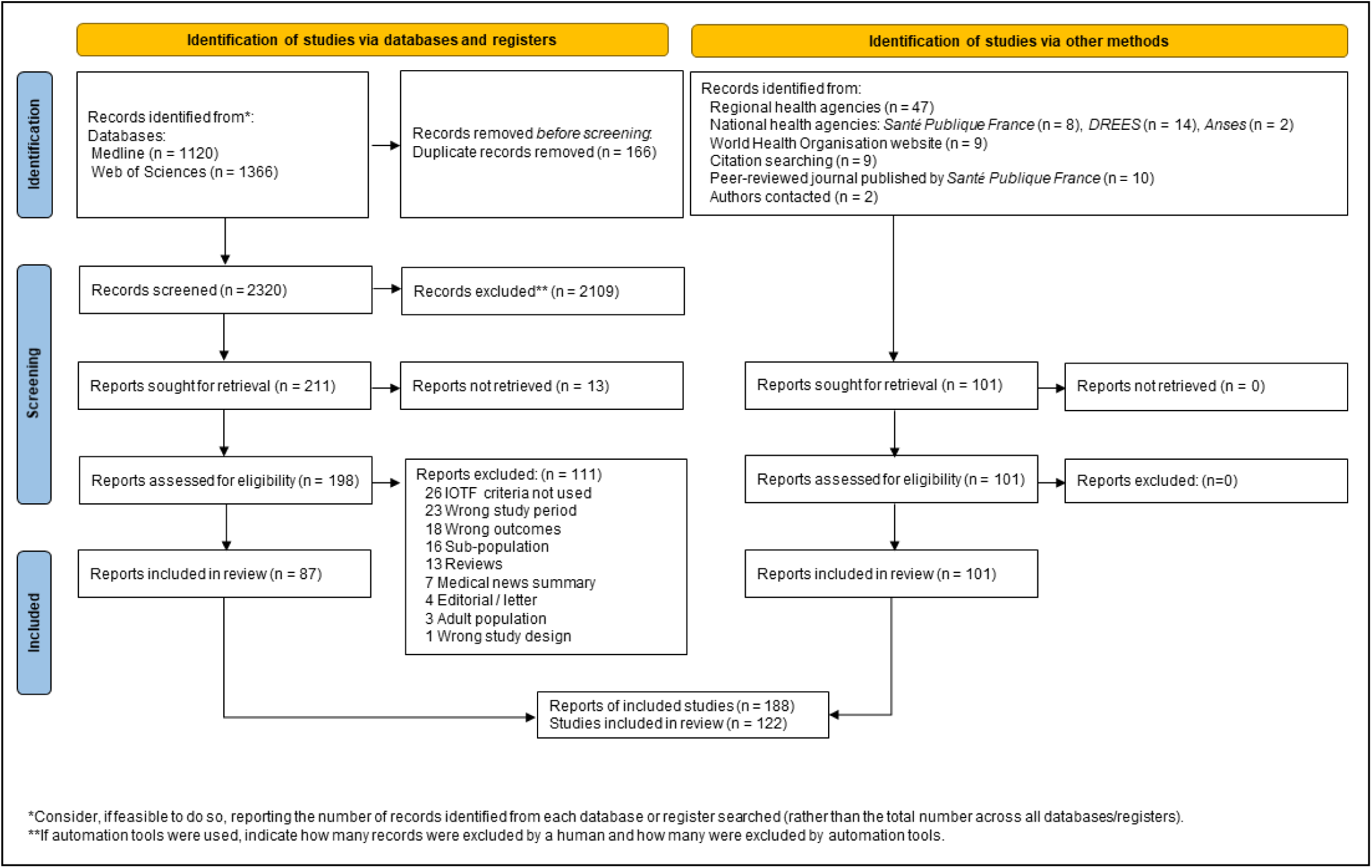
Study flow chart.

### Risk-of-bias assessments

Of the 122 studies, 17 (14%) were considered at unclear risk of bias and 77 (63%) at high risk of bias (**Supplemental Table 6**), mainly due to a lack of representative sample of the general population (n=41, 34%), high refusal or attrition rates (n=28, 23%) ranging from 45% to 91%, or the use of non-standardised weight measurement methods (n=23, 19%), with ≥ 2 reasons in 22% (n=27) of studies. (**Figure 2**). Of the 28 (23%) studies considered at low risk of bias, 18 were nationwide and included 143,655 children and adolescents aged 2 to 17 years (70,806 girls, 49%). No nationwide study at low risk of bias was available after 2017.

**Figure 2.**
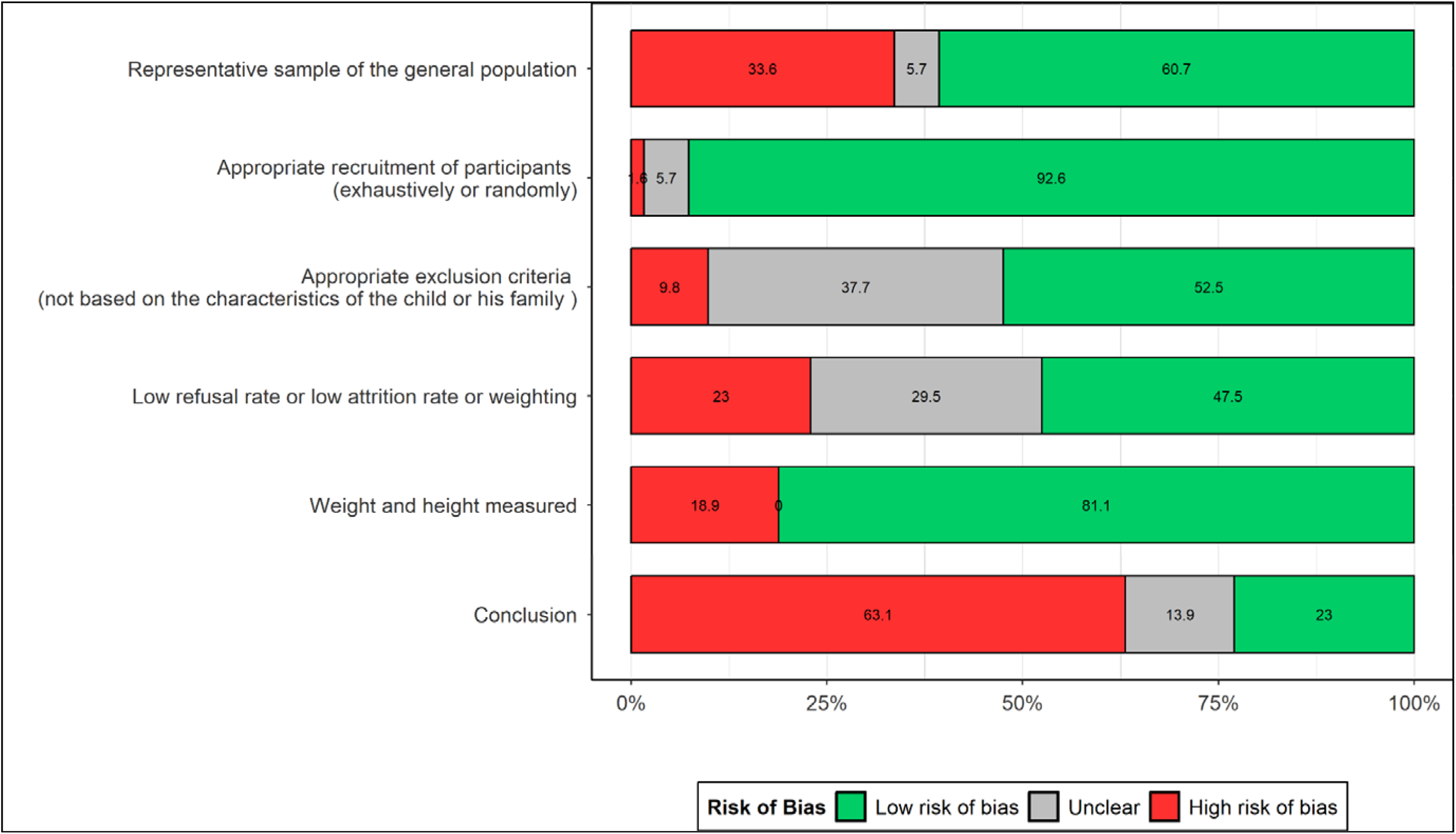
Risk-of-bias assessment.

### Summary prevalence of overweight and obesity

Among the 18 nationwide studies at low risk of bias, the prevalence of overweight (including obesity) ranged from 3.8% to 24.2% for girls and 3.9% to 21.4% for boys (**Figure 3**). Without stratification on sex, the summary estimate of the prevalence was 17.0% (95% CI 15.3-18.9) for age 2 to 17 years: 12.8% (11.5-14.0) for age 2 to 5 years, 18.9% (17.2-20.8) for age 6 to 11 years and 15.6% (12.6-19.0) for age 12 to 17 years. For girls, the summary estimate of the prevalence was 18.2% (16.5-20.0) for age 2 to 17 years: 14.3% (12.7-16.1) for age 2 to 5 years, 21.0% (19.5-22.4) for age 6 to 11 years, and 16.0% (12.6-19.7) for age 12 to 17 years. For boys, the summary estimate of the prevalence was 16.0% (14.2-17.9) for age 2 to 17 years: 11.2% (10.3-12.2) for age 2 to 5 years, 17.4% (15.3-19.5) for age 6 to 11 years and 15.6% (13.0-18.4) for age 12 to 17 years.

**Figure 3.**
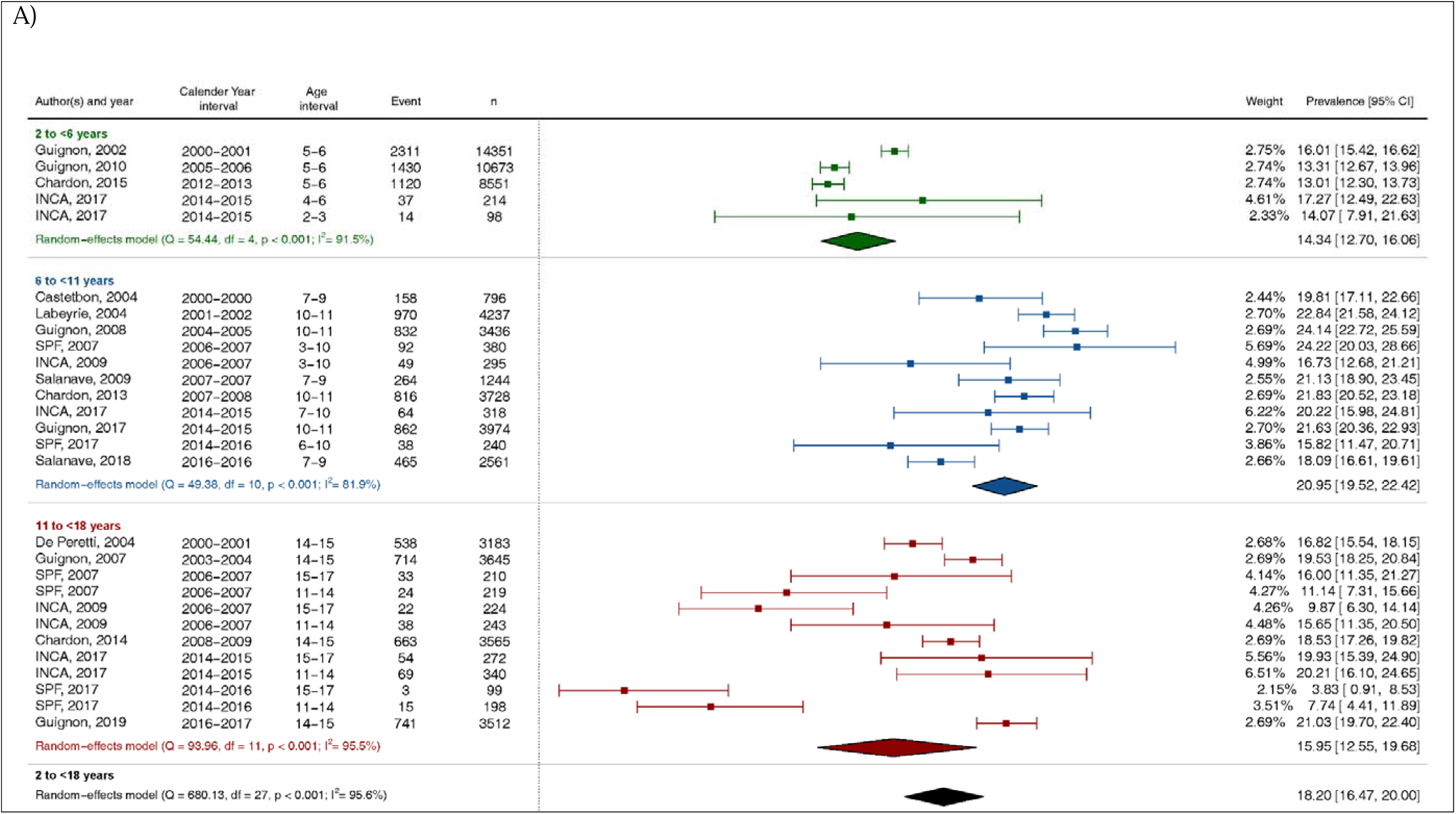
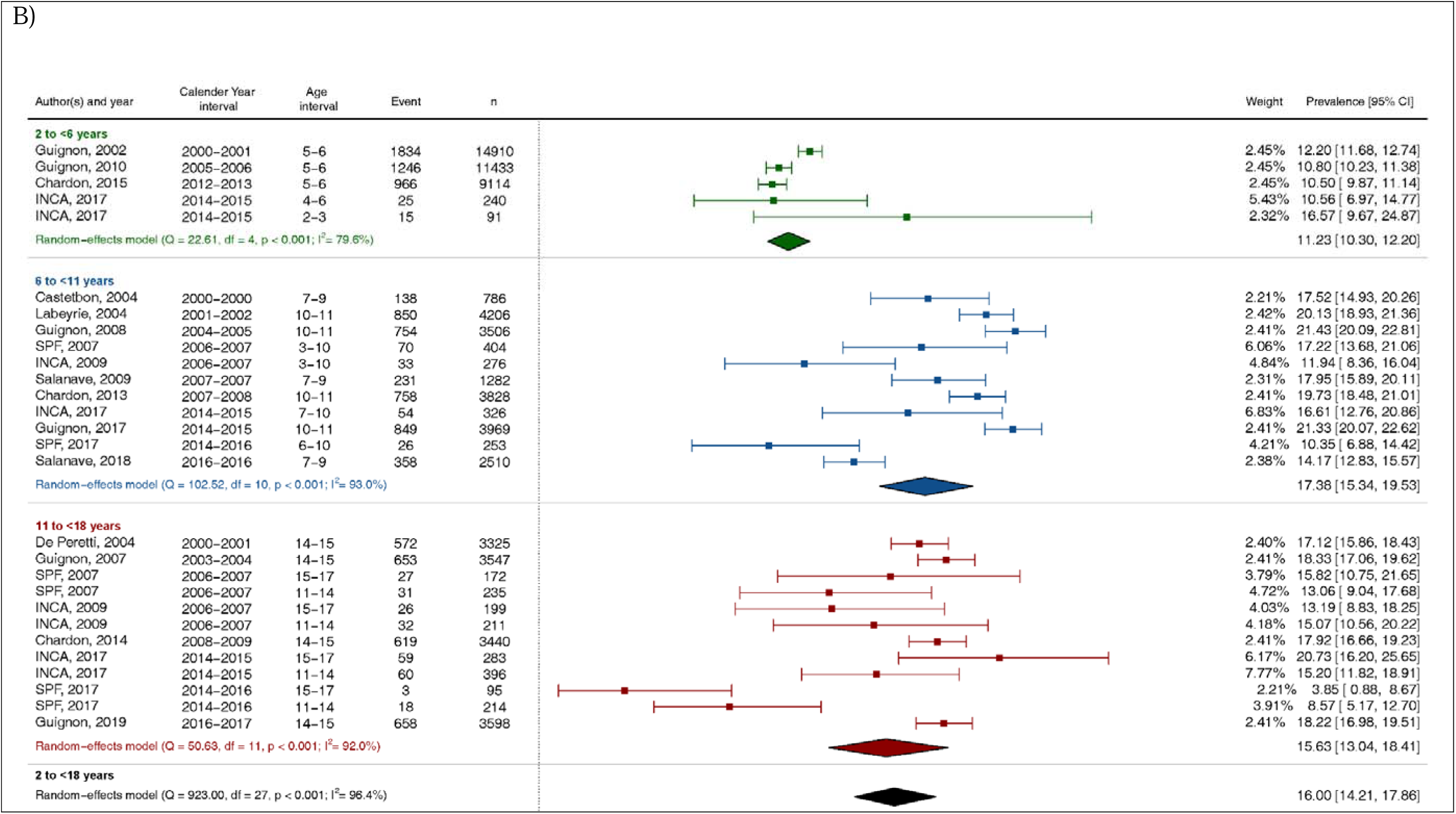
Meta-analysis results. Forest plot of 18 nationwide studies at low risk of bias reporting the prevalence of overweight (including obesity) in girls (A) and boys (B), globally and by age class.

Obesity prevalence varied from 1.3% to 8.3% for girls and 1.3% to 7.0% for boys (**Figure 4**). Without stratification on sex, the summary estimate of the prevalence of paediatric obesity was 4.0% (95% CI 3.6-4.5) for age 2 to 17 years: 3.3% (2.9-3.8) for age 2 to 5 years, 4.5% (4.2-4.8) for age 6 to 11 years and 3.6% (2.8-4.6) for age 12 to 17 years. For girls, the summary estimate was 4.3% (3.9-4.7) for age 2 to 17 years: 3.7% (3.0-4.5) for age 2 to 5 years, 4.9% (4.6-5.2) for age 6 to 11 years and 3.8% (3.0-4.8) for age 12 to 17 years. For boys, the summary estimate was 3.9% (3.4-4.3) for age 2 to 17 years: 3.0% (2.7-3.3) for age 2 to 5 years, 4.3% (3.9-4.7) for age 6 to 11 years and 3.7% (2.9-4.7) for age 12 to 17 years.

**Figure 4.**
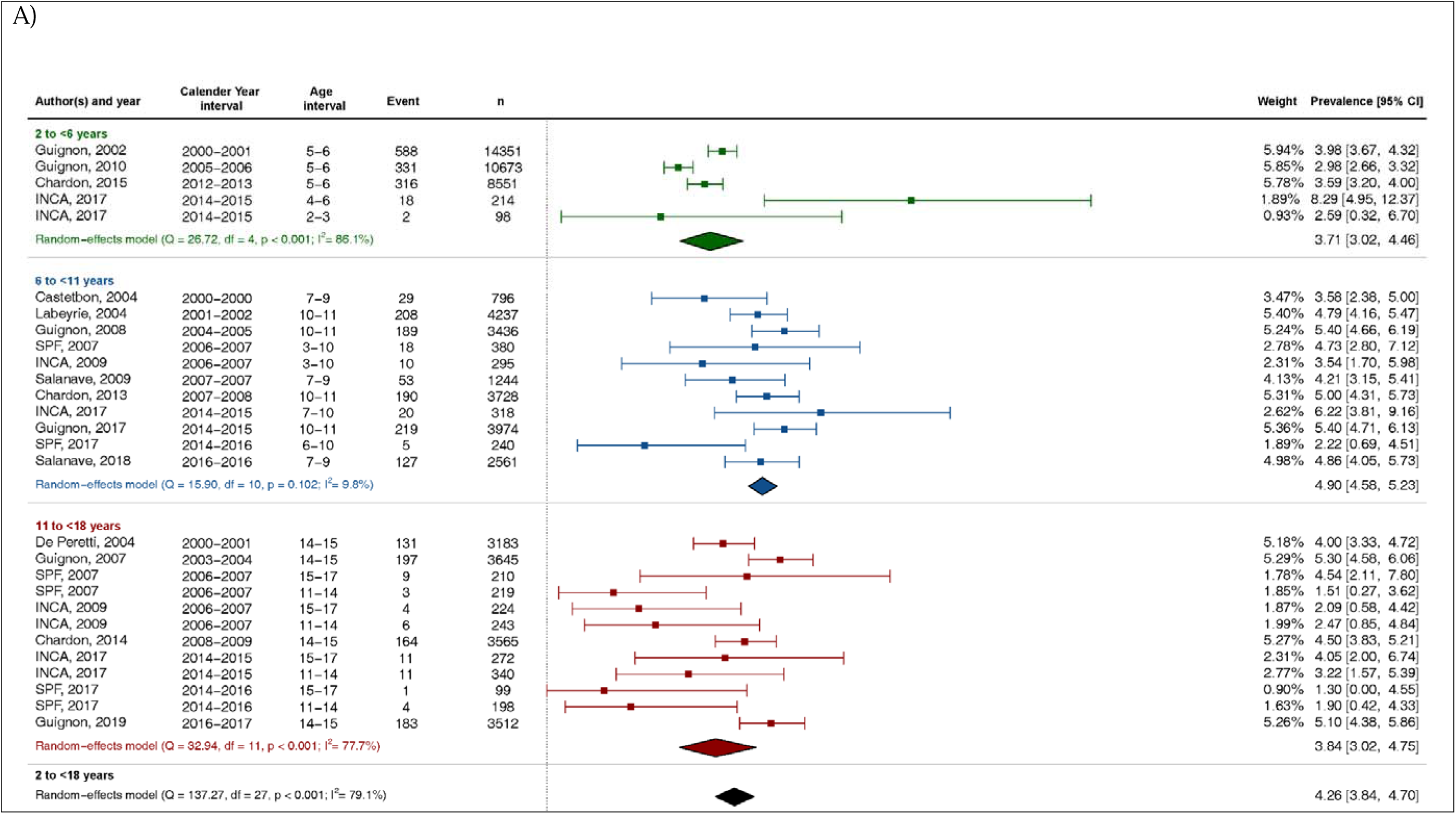
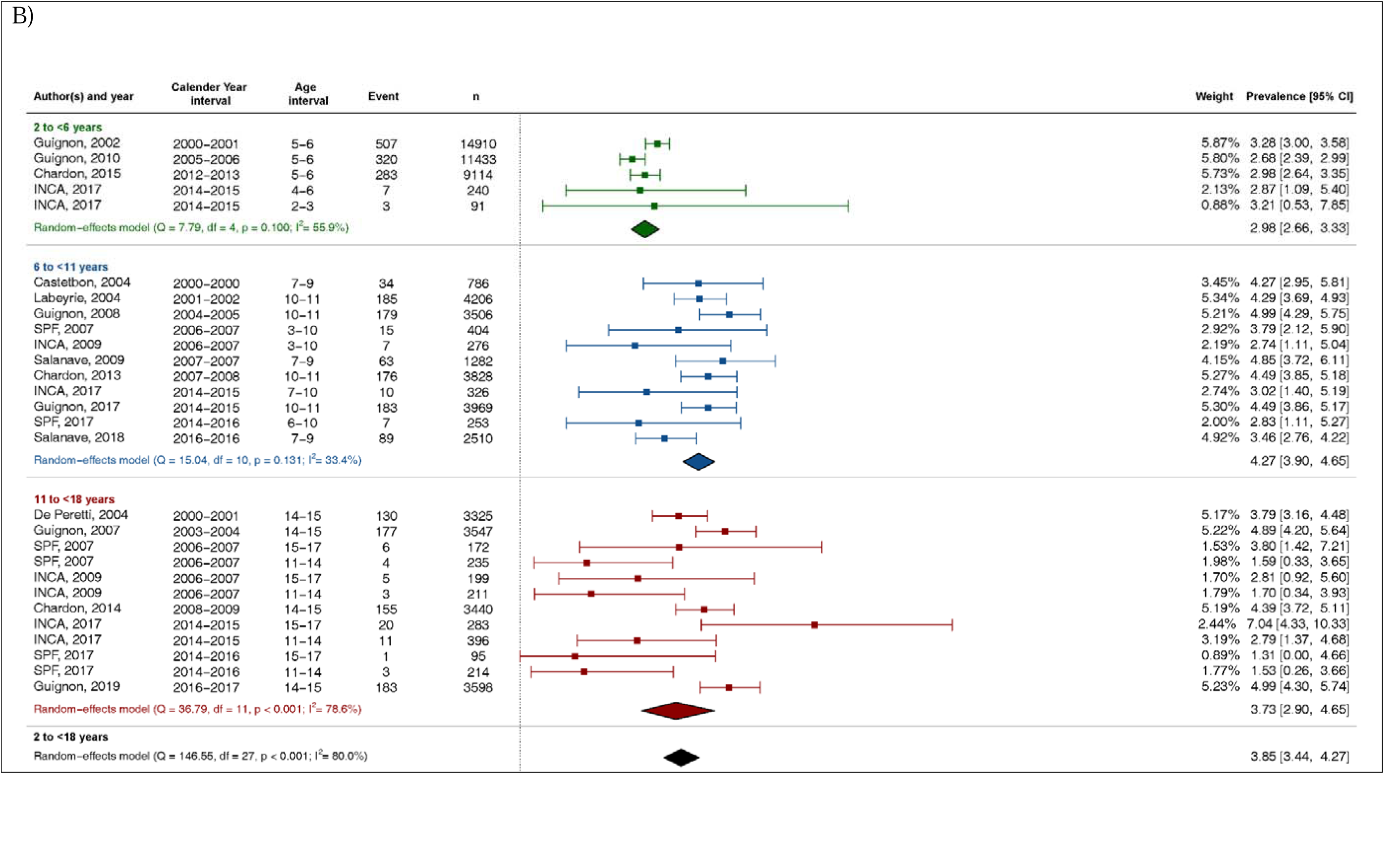
Meta-analysis results. Forest plot of 18 nationwide studies at low risk of bias reporting the prevalence of obesity in girls (A) and boys (B), globally and by age class.

Between-study heterogeneity was substantial for the prevalence of overweight (including obesity) and obesity for both sexes, globally and by age range, with I² >80%, except for obesity between age 6 and 11 years (I^2^ =9.6% and 33.4% for girls and boys, respectively).

### Temporal trends in the prevalence of overweight and obesity

The meta-regression for overweight (**Figure 5**) and obesity (**Figure 6**) revealed no significant interaction between calendar year midrange, age class, and sex. After adjustment for age class, we found no statistically significant temporal trends in the prevalence of overweight (including obesity) or obesity from 2000 to 2017 for girls (**Table 1**) or boys (**Table 2**).

**Figure 5.**
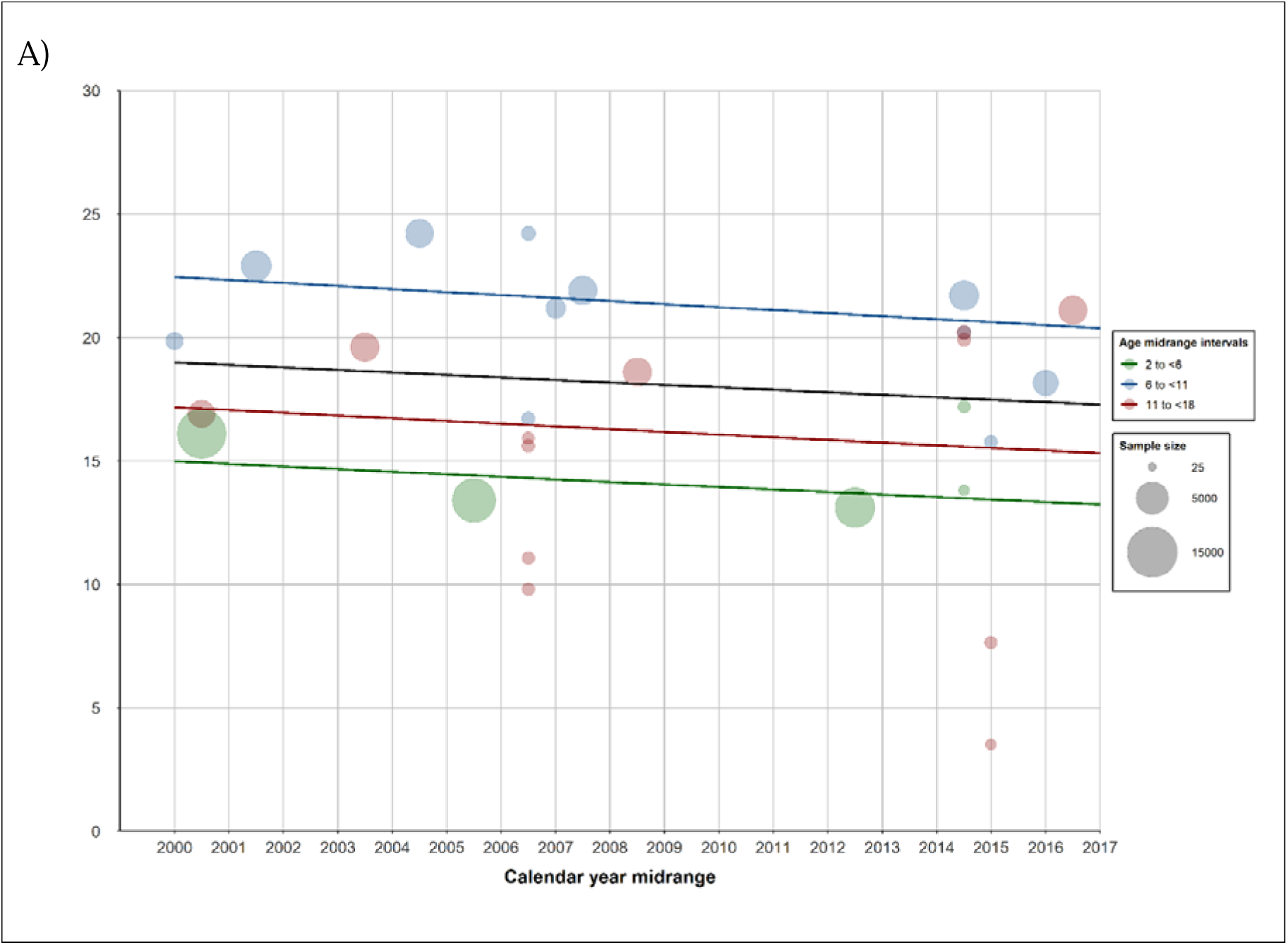
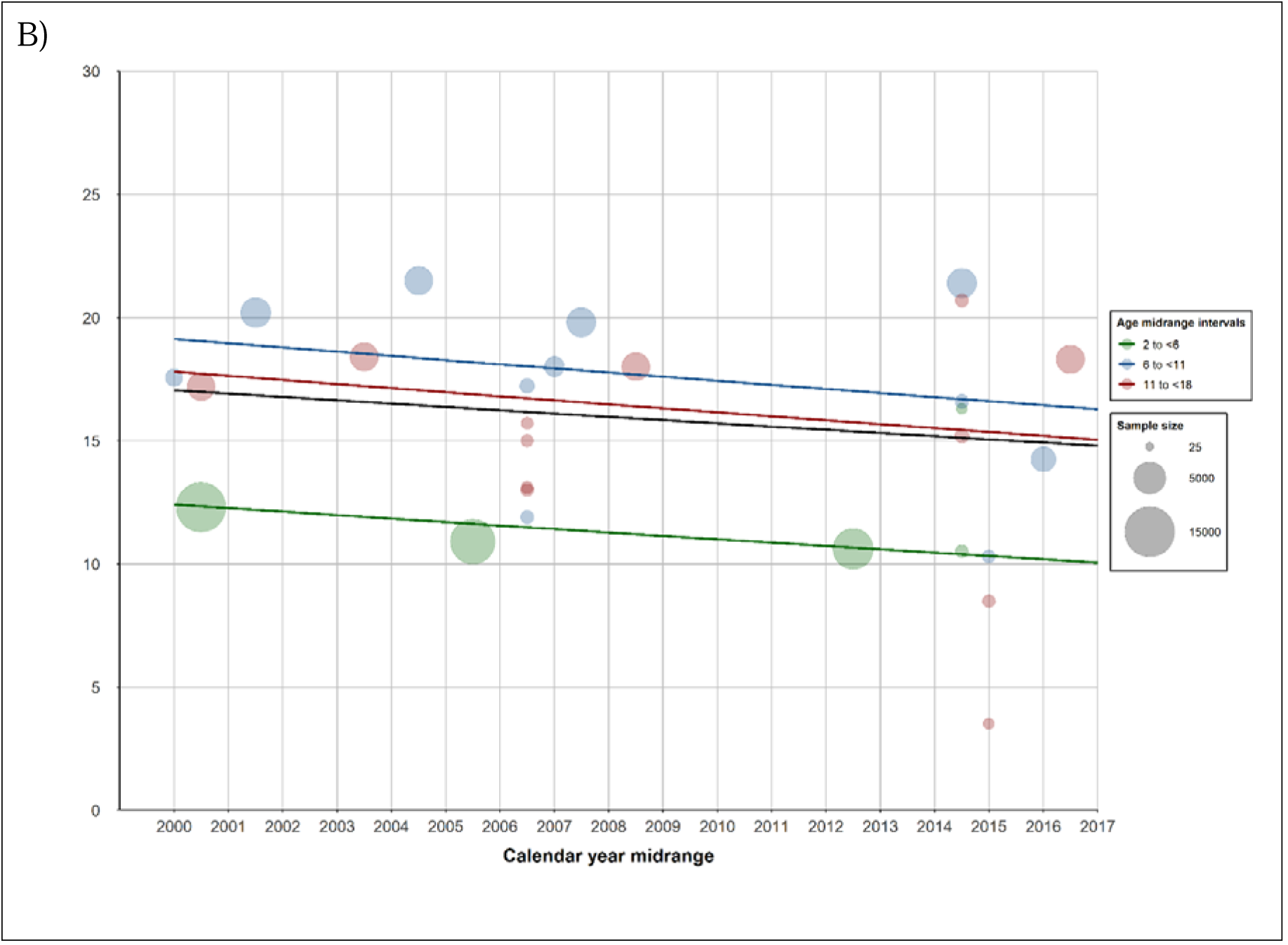
Measured (spots) and modelled (line) prevalence of overweight (including obesity) in girls (A) and boys (B) from 2000 to 2017 in France from 18 nationwide studies at low risk of bias, globally (black) and by age class 2 to <6 years (green), 6 to <11 years (blue), and 11 to <18 years (red). No study was available after 2017.

**Figure 6.**
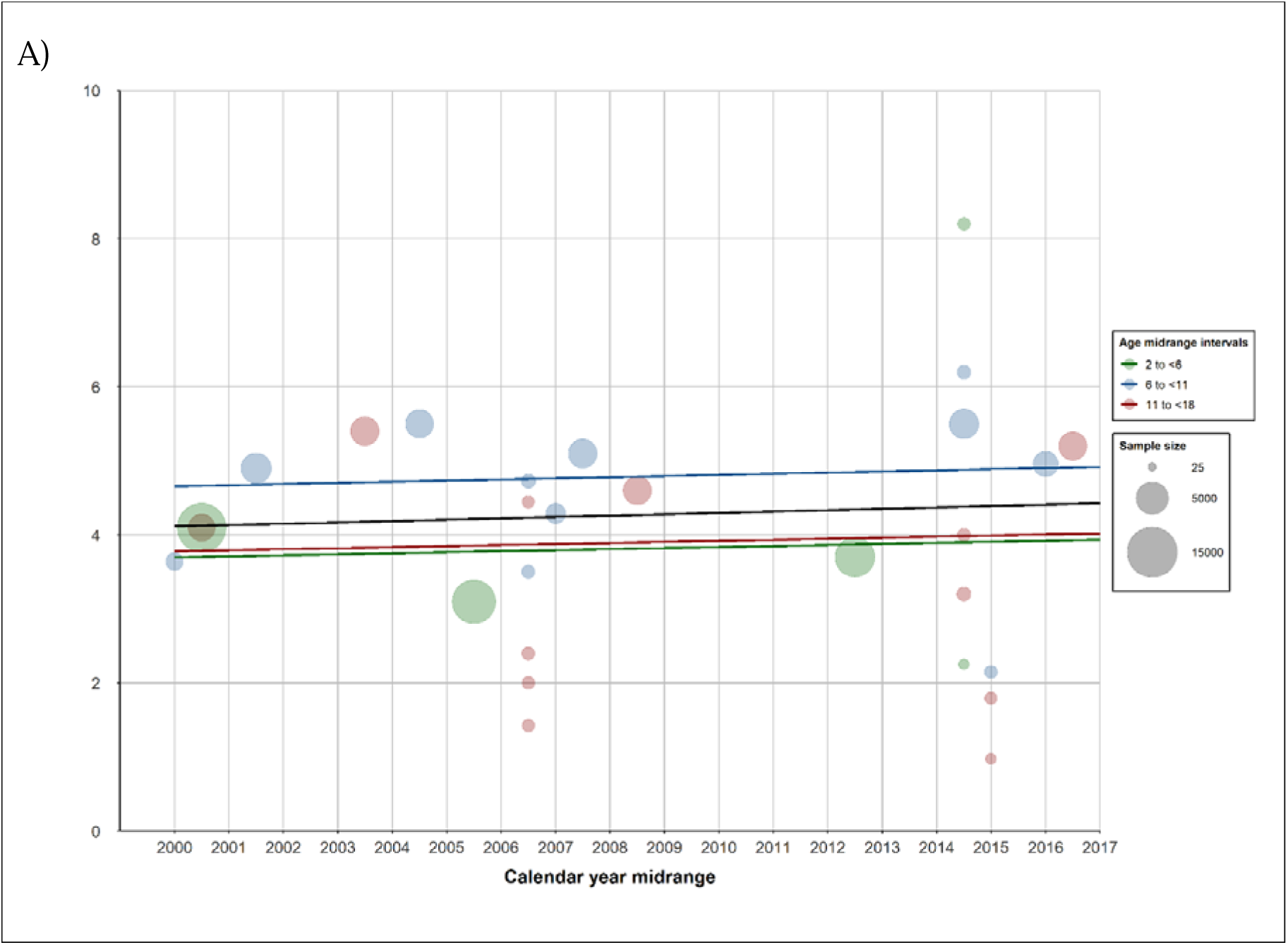
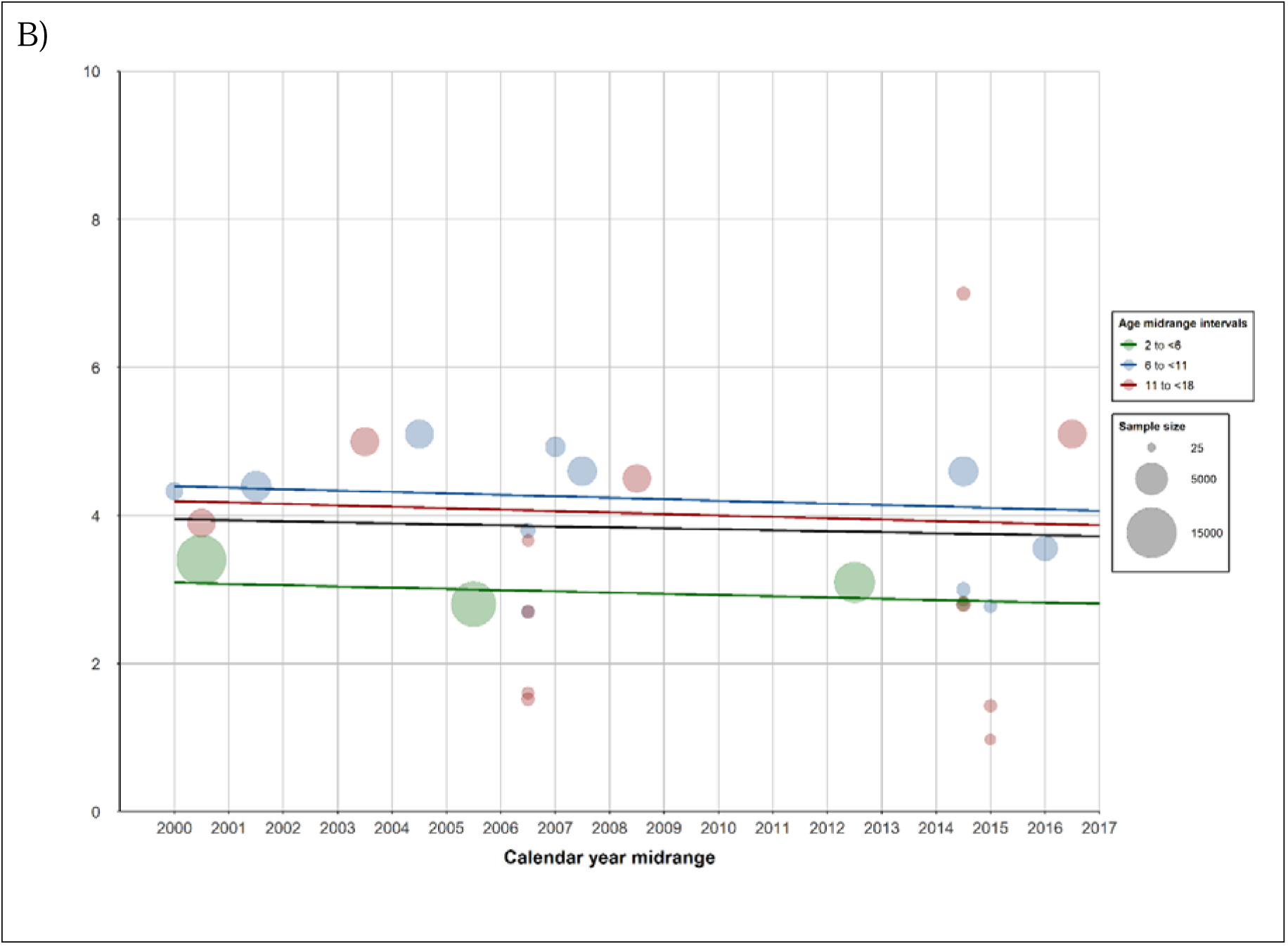
Measured (spots) and modelled (line) prevalence of obesity in girls (A) and boys (B) from 2000 to 2017 in France from 18 nationwide studies at low risk of bias, globally (black) and by age class 2 to <6 years (green), 6 to <11 years (blue), and 11 to <18 years (red). No nationwide study was available after 2017.

**Table 1.** Meta-regression results. Temporal trends in the prevalence of overweight (including obesity) and obesity in girls from 2000 to 2017 in France. No study was available after 2017.

|  | Nationwide low-risk studies (n=18, k=28) |  |  | All studies* |  |  |
| --- | --- | --- | --- | --- | --- | --- |
| | $\beta$ | [95% CI] | P-value | $\beta$ | [95% CI] | P-value |
| **Overweight (including obesity)** |  |  |  |  |  |  |
| Intercept | 14.99 | [12.01; 18.23] |  | 12.34 | [11.08; 13.66] |  |
| Calendar year midrange | -0.11** | [-0.31; 0.14] | 0.3857 | 0.11 | [0.04; 0.18] | **0.0020** |
| Age class |  |  |  |  |  |  |
| 2 to <6 |  | ref | < **0.0001** |  | ref | < **0.0001** |
| 6 to <11 | 7.46 | [3.78; 11.68] |  | 3.36 | [2.55; 4.24] |  |
| 11 to <18 | 2.18 | [-0.76; 5.81] |  | 3.64 | [2.65; 4.71] |  |
| **Obesity** |  |  |  |  |  |  |
| Intercept | 3.70 | [2.80; 4.71] |  | 2.60 | [2.15; 3.09] |  |
| Calendar year midrange | 0.01 | [-0.05; 0.10] | 0.7075 | 0.07 | [0.04; 0.11] | < **0.0001** |
| Age class |  |  |  |  |  |  |
| 2 to <6 |  | ref | 0.0735 |  | ref | **0.0035** |
| 6 to <11 | 0.96 | [-0.08; 2.35] |  | 0.51 | [0.17; 0.92] |  |
| 11 to <18 | 0.08 | [-0.78; 1.33] |  | 0.14 | [-0.20; 0.56] |  |

k: statistical unit. The $\beta$ estimates and their 95% CIs were back-transformed by using the Freeman-Tukey double arcsine.
\* For overweight (including obesity): n=97 studies with data available and k=215; for obesity: n=95 with data available and k=213.
\*\* Between 2000 and 2010, the prevalence of overweight (including obesity) decreased from 1.1 percentage points in children aged 2 to <6 years.

**Table 2.** Meta-regression results. Temporal trends in the prevalence of overweight (including obesity) and obesity in boys from 2000 to 2017 in France. No study was available after 2017.

|  | Nationwide low-risk of bias (n=18, k=28) |  |  | All studies* |  |  |
| --- | --- | --- | --- | --- | --- | --- |
| | $\beta$ | [95% CI] | P-value | $\beta$ | [95% CI] | P-value |
| **Overweight (including obesity)** |  |  |  |  |  |  |
| Intercept | 12.41 | [9.74; 15.35] |  | 10.60 | [9.52; 11.73] |  |
| Calendar year midrange | -0.14** | [-0.32; 0.08] | 0.1897 | -0.01 | [-0.07; 0.05] | 0.7545 |
| Age class |  |  |  |  |  |  |
| 2 to <6 |  | ref | < 0.0001 |  |  | <0 .0001 |
| 6 to <11 | 6.72 | [3.31; 10.70] |  | 3.47 | [2.70; 4.30] |  |
| 11 to <18 | 5.39 | [2.18; 9.21] |  | 7.24 | [6.11; 8.44] |  |
| **Obesity** |  |  |  |  |  |  |
| Intercept | 3.09 | [2.44; 3.82] |  | 1.96 | [1.56; 2.41] |  |
| Calendar year midrange | -0.02 | [-0.06; 0.04] | 0.5301 | 0.04 | [0.01; 0.07] | 0.0017 |
| Age class |  |  |  |  |  |  |
| 2 to <6 |  | ref | 0.0044 |  |  | < 0.0001 |
| 6 to <11 | 1.31 | [0.43; 2.40] |  | 0.70 | [ 0.36; 1.10] |  |
| 11 to <18 | 1.10 | [0.23; 2.21] |  | 1.35 | [0.87; 1.91] |  |

k: statistical unit. The $\beta$ estimates and their 95% CIs were back-transformed by using the Freeman-Tukey double arcsine.
\* For overweight (including obesity): n=97 studies with data available and k=215; for obesity: n=95 with data available and k=213.
\*\* Between 2000 and 2010, the prevalence of overweight (including obesity) decreased from 1.4 percentage points in children aged 2 to <6 years.

### Sensitivity analysis

Of the 122 included studies, 97 and 95 reported complete data for overweight and obesity, respectively. The studies included 669,434 (331,824 girls, 50%) and 638,339 (316,601 girls, 50%) children and adolescents aged 2 to 17 years, respectively. The summary estimate of the prevalence of overweight (including obesity) was 16.1% (14.9-17.3) for girls and 14.9% (13.7-16.1) for boys. The summary estimate of the prevalence of obesity was 3.5% (3.2-3.9) for girls and 3.3% (2.9-3.7) for boys. We found significant positive temporal trends in the prevalence of overweight (including obesity) and obesity in girls and boys, except for overweight in boys (**Tables 1** and **2**).

The use of logit transformation in meta-analyses gave similar summary estimates (**Supplemental Figures 2** and **3**) and trends (**Supplemental Tables 7** and **8**) for overweight and obesity for both sexes.

## DISCUSSION

### Main findings and interpretation

This systematic review provided a comprehensive overview of 122 studies reporting the prevalence of overweight and obesity, defined by BMI and IOTF thresholds, in more than half a million children and adolescents aged 2 to 17 years across France from 2000 to 2024. The estimated prevalence of overweight (including obesity) was 18.2% for girls and 16.0% for boys, whereas that for obesity alone was 4.3% for girls and 3.9% for boys.

These results can be compared with those of two previous international meta-analyses that examined prevalence across several countries, including France.^3, 16^ For the years 2000 to 2013, Ng *et al* reported a higher prevalence of overweight (including obesity) in France (girls: 16.7–16.0% and boys: 21.2–19.9%;) and obesity (girls: 4.4–4.7% and boys: 5.5–5.8%).^16^ In 2021, a similar prevalence was reported.^29^ Zhang *et al* reported a lower prevalence of overweight (including obesity) (13.0% vs. 17.0%) but a similar prevalence of obesity (3.9% vs. 4.0%) between 2020 and 2023.^3^ The difference with our results may be explained by the exclusion of potential eligible individuals in previous analyses, the inclusion in the Zhang *et al* meta-analysis of studies that did not use the IOTF definition,^30–32^ and/or the lack of risk-of-bias assessment, which may have led to the inclusion of biased studies.

We found higher overweight (including obesity) prevalence in children aged 6 to 11 years (18.9%; girls 21.0% and boys 17.4%) than 2 to 5 years (12.8%; girls 14.3% and boys 11.2%) and 12 to 17 years (15.6%; girls 16% and boys 15.6%), particularly girls. These age- and sex-specific prevalence patterns, in particular between 6-11 and 12-17 years, may be explained by hormonal changes during puberty.^33^ This earlier hormonal change in girls may help explain the higher prevalence of overweight in girls than boys aged 6 to 11 years. Another possible explanation is lower levels of physical activity in girls, which may contribute to the higher prevalence of overweight in girls aged 11 to 16 years.^34^

We found no significant temporal trend in prevalence of overweight (including obesity) and obesity in children and adolescents in France from 2000 to 2017 after adjustment for age class. Such stabilization of paediatric overweight and obesity over the past 2 decades was previously observed in some European countries (including France) as well as in other high-income countries, with different prevalence levels.^1, 2, 17, 35^ Increasing public awareness of obesity as major health issue has led to the implementation of prevention strategies promoting daily physical activity and healthy eating, which may have played a role in this stabilization^18,36^ However, some studies suggested that the prevalence of overweight and obesity has increased since the 2000s among children and adolescents from disadvantaged families, in contrast to the stagnation or decline observed among their more advantaged peers.^37–39^ Importantly, prevention strategies focusing on individual behaviour change seem less effective in socially disadvantaged than advantaged populations, potentially contributing to the widening of health disparities.^40^

We did not identify any studies at low risk of bias reporting paediatric overweight and obesity prevalence in France after 2017. This gap is particularly striking for a country with the 7^th^ largest economy in the world. As a result, we cannot evaluate the potential impact of the COVID-19 pandemic on overweight and obesity prevalence in France. In other countries, school closures, reduced access to physical activity, increased screen time, or psychological stress, related to the pandemic and lockdowns, may have contributed to an increase in prevalence.^41–47^

Most studies (77%, n=94/122) were at high or unknown risk of bias, primarily due to poor design or reporting. The three main causes of bias were lack of representativeness, high refusal or attrition rates, or the use of non-standardised weight measurement methods. Meta-analyses of prevalence data require careful assessment of the risk of bias, and using data from studies at low risk of bias is crucial for obtaining reliable summary estimates.^48^ In the sensitivity analyses including all studies regardless of risk of bias, the prevalence estimates were lower (e.g., 18.2% vs. 16.1% for overweight in girls), and we found a significant positive temporal trend in overweight, except for boys. These results highlight the need for a careful analysis of the risk of bias in prevalence meta-analyses.

### Implications

The results of this present study highlight the need for repeated high-quality studies to regularly monitor trends in the prevalence of obesity and overweight.^39, 40^ National surveillance systems have been established in several countries, including Australia, England, Canada, and the United States, with varying objectives, methodologies, or data collection approaches.^49^ International obesity monitoring has also been proposed through national representative school surveys (2009-2022) conducted at three different ages (11, 13, and 15 years) as well as representative nationwide studies (2007-2020) in children aged 6 to 9 years in approximately 40 participating countries, including France.^50–55^ However, these monitoring systems have a limited age range coverage and operate irregularly over time. Therefore, the challenge would be to develop a regular and rigorous monitoring system covering the entire population from birth to age 17, ensuring adequate representation of the paediatric population from overseas departments and regions as well as those from socially disadvantaged families.^11, 56–59^ To meet this goal, a medical observatory gathering growth data routinely collected in clinical practice could be part of the solution, as demonstrated by publication of the new French growth charts.^60, 61^ Once established, this comprehensive mapping of paediatric overweight or obesity prevalence will support the implementation of early effective strategies to prevent paediatric obesity. It would target structural changes to the environment to reduce health inequalities,^62^ grounded in Marmot’s proportionate universalism approach.^63,64^

### Strengths and limitations

This systematic review is based on an exhaustive search of published studies of French data, including data from international, regional, and national health agency websites. Although this systematic review and meta-analysis followed a rigorous methodological guideline for reporting studies, some limitations should be acknowledged. Some are common to meta-analyses (e.g., selection bias and limited availability of complete information from study reports), but others are specific to our study. First, the meta-analysis based on only 18 studies with low risk of bias, corresponding to 28 statistical units, may have led to insufficient power to obtain accurate estimates of summary prevalence and to study prevalence trends. Studies that used definitions other than the IOTF standard were excluded from the systematic review, which led to a reduced number of studies in the meta-analysis. However, the number of such studies was few because the IOTF definition is currently the recommended standard in France. Second, we were not able to analyse the source of variation in prevalence, notably geographic area; however, because of the limited number of non-nationwide studies at low risk of bias, estimating this prevalence within French regions, especially in overseas departments and regions, is difficult.

## CONCLUSION

We found no significant increase in the paediatric prevalence of overweight and obesity in France since 2000. This conclusion is based on a small number of studies with high methodological quality that reported prevalence only up to 2017, which precludes an investigation of the potential impact of the COVID-19 lockdown. Of note, the inclusion of studies regardless of risk of bias led to different conclusions. Regular, high-quality nationwide studies are needed to rigorously monitor trends in paediatric overweight and obesity and to assess the effectiveness of related policies.

## Data Availability

All relevant data are within the paper.

## REGISTRATION AND PROTOCOL

The protocol of this review was registered (no CRD42022352404; https://www.crd.york.ac.uk/prospero/display_record.php?RecordID=352404).

## CONTRIBUTORS (CRediT)

**Marianne Jacques.** Investigation; Data curation; Formal analysis; Writing - Original Draft; Visualization

**Martin Chalumeau.** Conceptualization; Supervision; Project administration; Writing - Review & Editing; Funding acquisition

**Camille Le Gal.** Investigation; Data curation; Writing - Original Draft

**Benoit Salanave.** Investigation; Writing - Review & Editing

**Bruno Frandji.** Writing - Review & Editing

**Marie-Aline Charles.** Writing - Review & Editing

**Sandrine Lioret.** Investigation; Writing - Review & Editing

**Barbara Heude.** Conceptualization; Supervision; Project administration; Writing - Review & Editing

**Pauline Scherdel.** Conceptualization; Methodology; Software; Visualization; Supervision; Project administration; Writing - Review & Editing

## DATA AVAILABILITY STATEMENT

All relevant data are within the paper.

## ABBREVIATIONS

BMI: body mass index
IOTF: International Obesity Task Force
WHO: World Health Organisation

