## Appendices for "Trends in the prevalence of overweight and obesity in children and adolescents in France in the 21^st^ century: a systematic review and meta-analysis"

### SUPPLEMENTAL MATERIALS

#### Supplemental Table 1 Preferred reporting items for systematic reviews and meta-analyses (PRISMA) statement.^1^

| **Section and Topic** | **Item #** | **Checklist item** | **Location where item is reported** |
| --- | --- | --- | --- |
| **TITLE** | | |  |
| Title | 1 | Identify the report as a systematic review. | 1 |
| **ABSTRACT** | | |  |
| Abstract | 2 | See the PRISMA 2020 for Abstracts checklist. | 8 |
| **INTRODUCTION** | | |  |
| Rationale | 3 | Describe the rationale for the review in the context of existing knowledge. | 10 |
| Objectives | 4 | Provide an explicit statement of the objective(s) or question(s) the review addresses. | 11 |
| **METHODS** | | |  |
| Eligibility criteria | 5 | Specify the inclusion and exclusion criteria for the review and how studies were grouped for the syntheses. | 11-12 |
| Information sources | 6 | Specify all databases, registers, websites, organisations, reference lists and other sources searched or consulted to identify studies. Specify the date when each source was last searched or consulted. | 11-12 |
| Search strategy | 7 | Present the full search strategies for all databases, registers and websites, including any filters and limits used. | 11-12 |
| Selection process | 8 | Specify the methods used to decide whether a study met the inclusion criteria of the review, including how many reviewers screened each record and each report retrieved, whether they worked independently, and if applicable, details of automation tools used in the process. | 11-12 |
| Data collection process | 9 | Specify the methods used to collect data from reports, including how many reviewers collected data from each report, whether they worked independently, any processes for obtaining or confirming data from study investigators, and if applicable, details of automation tools used in the process. | 11 |
| Data items | 10a | List and define all outcomes for which data were sought. Specify whether all results that were compatible with each outcome domain in each study were sought (e.g. for all measures, time points, analyses), and if not, the methods used to decide which results to collect. | 12 |
|  | 10b | List and define all other variables for which data were sought (e.g. participant and intervention characteristics, funding sources). Describe any assumptions made about any missing or unclear information. | 12 |
| Study risk of bias assessment | 11 | Specify the methods used to assess risk of bias in the included studies, including details of the tool(s) used, how many reviewers assessed each study and whether they worked independently, and if applicable, details of automation tools used in the process. | 13 |
| Effect measures | 12 | Specify for each outcome the effect measure(s) (e.g. risk ratio, mean difference) used in the synthesis or presentation of results. | 13 |
| Synthesis methods | 13a | Describe the processes used to decide which studies were eligible for each synthesis (e.g. tabulating the study intervention characteristics and comparing against the planned groups for each synthesis (item #5)). | 13-14 |
|  | 13b | Describe any methods required to prepare the data for presentation or synthesis, such as handling of missing summary statistics, or data conversions. |  |
|  | 13c | Describe any methods used to tabulate or visually display results of individual studies and syntheses. | 13-14 |
|  | 13d | Describe any methods used to synthesize results and provide a rationale for the choice(s). If meta-analysis was performed, describe the model(s), method(s) to identify the presence and extent of statistical heterogeneity, and software package(s) used. | 13-14 |
|  | 13e | Describe any methods used to explore possible causes of heterogeneity among study results (e.g. subgroup analysis, meta-regression). |  |
|  | 13f | Describe any sensitivity analyses conducted to assess robustness of the synthesized results. | 13-14 |
| Reporting bias assessment | 14 | Describe any methods used to assess risk of bias due to missing results in a synthesis (arising from reporting biases). | NA |
| Certainty assessment | 15 | Describe any methods used to assess certainty (or confidence) in the body of evidence for an outcome. | NA |
| **RESULTS** | | |  |
| Study selection | 16a | Describe the results of the search and selection process, from the number of records identified in the search to the number of studies included in the review, ideally using a flow diagram. | 14-15 |
|  | 16b | Cite studies that might appear to meet the inclusion criteria, but which were excluded, and explain why they were excluded. | 14-15 |
| Study characteristics | 17 | Cite each included study and present its characteristics. | 14-15 |
| Risk of bias in studies | 18 | Present assessments of risk of bias for each included study. | 15 |
| Results of individual studies | 19 | For all outcomes, present, for each study: (a) summary statistics for each group (where appropriate) and (b) an effect estimate and its precision (e.g. confidence/credible interval), ideally using structured tables or plots. | 15-16 |
| Results of syntheses | 20a | For each synthesis, briefly summarise the characteristics and risk of bias among contributing studies. |  |
|  | 20b | Present results of all statistical syntheses conducted. If meta-analysis was done, present for each the summary estimate and its precision (e.g. confidence/credible interval) and measures of statistical heterogeneity. If comparing groups, describe the direction of the effect. | 16 |
|  | 20c | Present results of all investigations of possible causes of heterogeneity among study results. | 16 |
|  | 20d | Present results of all sensitivity analyses conducted to assess the robustness of the synthesized results. | 17 |
| Reporting biases | 21 | Present assessments of risk of bias due to missing results (arising from reporting biases) for each synthesis assessed. | NA |
| Certainty of evidence | 22 | Present assessments of certainty (or confidence) in the body of evidence for each outcome assessed. | NA |
| **DISCUSSION** | | |  |
| Discussion | 23a | Provide a general interpretation of the results in the context of other evidence. | 17-19 |
|  | 23b | Discuss any limitations of the evidence included in the review. | 20-21 |
|  | 23c | Discuss any limitations of the review processes used. | 20-21 |
|  | 23d | Discuss implications of the results for practice, policy, and future research. | 19-21 |
| **OTHER INFORMATION** | | |  |
| Registration and protocol | 24a | Provide registration information for the review, including register name and registration number, or state that the review was not registered. | 21 |
|  | 24b | Indicate where the review protocol can be accessed, or state that a protocol was not prepared. | NA |
|  | 24c | Describe and explain any amendments to information provided at registration or in the protocol. | NA |
| Support | 25 | Describe sources of financial or non-financial support for the review, and the role of the funders or sponsors in the review. | 22 |
| Competing interests | 26 | Declare any competing interests of review authors. | 22 |
| Availability of data, code and other materials | 27 | Report which of the following are publicly available and where they can be found: template data collection forms; data extracted from included studies; data used for all analyses; analytic code; any other materials used in the review. | 22 |

#### Supplemental Table 2 Search box.

**PubMed**

1. ("child*"[Title/Abstract] OR "child"[MeSH Terms] OR "child, preschool"[MeSH Terms] OR "adolescent"[MeSH Terms])

2. ("obesity"[MeSH Terms] OR "obes*"[Title/Abstract] OR "overweight"[MeSH Terms] OR "overweight*"[Title/Abstract] OR "body mass index"[MeSH Terms] OR "body mass index"[Title/Abstract] OR "body composition"[MeSH Terms] OR "body weight"[MeSH Terms] OR "weight"[Title/Abstract])

3. ("France"[MeSH Terms] OR "France"[Title/Abstract] OR "French Overseas"[Title/Abstract])

4. (2000/01/01:2024/12/31 [Date - Publication]) NOT (("preterm"[Title/Abstract] OR "prema*"[Title/Abstract] OR "respiratory"[Title/Abstract] OR "surgery"[Title/Abstract] OR "apnea"[Title/Abstract]))

5. 1 AND 2 AND 3 AND 4

**Web of Science**

1. (KP=(child)) OR (KP=(adolescent))

2. (TS=(obes*)) OR (TS=(overweight*)) OR (TS=(“body mass index”)) OR (TS=(“body mass index”))

3. (CU=(France)) OR (TS=(France))

4. (PY=(2000-2024))

5. (TS=(preterm)) OR (TS=(prema*)) OR (TS=(respiratory)) OR (TS=(surgery)) OR (TS=(apnea))

6. 1 AND 2 AND 3 AND 4 NOT 5

7. Indexes: SCI-EXPANDED, SSCI, A&HCI, CPCI-S, CPCI-SSH, BKCI-S, BKCI-SSH, ESCI, CCR-EXPANDED, IC

#### Supplemental Table 3 Lists of regional, national, and international health agency websites.

| **Name of health agencies** | **Website** |
| --- | --- |
| *Agence Nationale Sécurité Sanitaire Alimentaire Nationale (Anses)* | [https://www.anses.fr/fr/content/les-%C3%A9tudes-inca](https://www.anses.fr/fr/content/les-études-inca) |
| *Direction de la Recherche, des Études, de l'Évaluation et des Statistiques (DRESS)* | <https://drees.solidarites-sante.gouv.fr/sources-outils-et-enquetes/les-enquetes-nationales-sur-la-sante-des-enfants-et-adolescents> |
| *Health Behavior in School-aged Children (HBSC)* | <https://www.who.int/europe/initiatives/health-behaviour-in-school-aged-children-(hbsc)-study>  <https://hbsc.org/> |
| *Observatoire Régional de la Santé (ORS)* | <https://www.fnors.org/les-publications/>  [www.orsbretagne.fr](http://www.orsbretagne.fr/)  [www.or2s.fr](http://www.or2s.fr/)  <https://orscreainormandie.org/>  <https://www.orsbfc.org/>  [www.orscentre.org](http://www.orscentre.org/)  <https://www.orspaysdelaloire.com/>  <https://www.ors-idf.org/>  <https://ors-ge.org/>  <http://www.ors-auvergne-rhone-alpes.org/>  <https://www.ors-na.org/>  <http://www.orspaca.org/>  <https://creaiors-occitanie.fr/>  <https://www.ors.corsica/>  <https://orsag.fr/>  <https://www.ors-martinique.org/>  <https://www.ors-guyane.org/>  <https://www.ors-reunion.fr/>  <https://ors-mayotte.org/> |
| *Santé Publique France (SPF)* | <https://www.santepubliquefrance.fr/publications> |

#### Supplemental Table 4 Customised questions to assess the risk of bias.

| **Questions** | | **Response** | |
| --- | --- | --- | --- |
|  |  | **Yes** | **No** |
| **Selection bias** |  | | |
|  | **1. Did the sampling plan allow for providing results representative of the population of the age group and the area studied, and was the sampling appropriately accounted for in the statistical analyses?** | The source population selected (or if applicable clusters) allowed for the inclusion of children and adolescents representative of the age group and the area studied, and was randomly selected or an appropriate statistical management of the sample selection strategy was used.  *Examples:*   - *The number of public and private schools included is proportional to the number of schools of each status in the study area* - *If the sampling design was stratified on certain characteristics (type of school etc.), the estimated prevalence accounted for these variables.* - *Although not all schools were surveyed, eligible schools were randomly selected.* | The general population belonged to a sub-population that had specific characteristics. There was a lack of appropriate statistical management of the sample selection strategy.  *Examples:*   - *The sample unit chosen is only private elementary schools (or only public ones)* - *Inclusion based on a medical follow-up with a particular professional category (maternal and child protection services, paediatrician)* - *Inclusion based on the existence of a specific pathology* |
|  | **2. Were study participants recruited appropriately among the source population?** | Eligible children and adolescents were recruited exhaustively or randomly from the selected source population. | The sample selection of eligible children and adolescents was not exhaustive or at random. |
|  | **3.** **Did the authors avoid inappropriate exclusion?** | No exclusion was based on child or family characteristics. | Some eligible children and adolescents were excluded based on child or family characteristics.  *Example: Specific exclusion of children and adolescents* *with a medical contraindication to the practice of a sport, chronic diseases etc.* |
|  | **4.** **Initial refusal to participate was low (arbitrarily defined below the first quartile of reported refusal rates of all included studies) or appropriately addressed, and in case of longitudinal studies, attrition was appropriately weighted** | Low initial refusal/lost to follow-up rates (longitudinal studies).  High participation refusal/attrition appropriately addressed by weighting if the characteristics of the analysed and the non-analysed children and adolescents differed. | No sample correction in case of high rates of participation refusal or attrition. |
| **Classification bias** |  | | |
|  | **5. Has the condition been accurately measured?** | Standardised measurement of anthropometric data, with individuals wearing light clothes and no shoes, using a scale and height gauge with low measuring inaccuracy.  *Example: Light clothes, no shoes* | Inappropriate measurement method.  *Examples:*   - *Parents reported weight or height* - *Children and adolescents* *wore their clothes/shoes* - *The error of the scale was more than +/- 100 g, or the error of the height gauge was more than 1 cm* |
| **Risk of bias** | - **High** if ≥1 ‘No’ for one of the questions - **Unclear** if ≥1 ‘Unclear’ for one of the questions and ‘Yes’ for the others - **Low** if ‘Yes’ to all questions | | |

#### Supplemental Table 5 Summary of 122 unique studies included.

|  | **Author, publication year** | **Type  publication** | **Survey  name** | **Calendar year interval** | **Study design** | **Location level** | **Location in France** | **Age interval  (year)** |
| --- | --- | --- | --- | --- | --- | --- | --- | --- |
| **1** | Aisne, 2003-04,^2^ | R |  | 2003-2004 | Cross sectional | Departmental | Aisne | 10-16 |
| **2** | Alsace, 2014-15,^3^ Alsace, 2014-16,^4^  Alsace, 2014-17,^5^ | R R R |  | 2014-2017 | Cross sectional | Regional | Alsace | 11-12 |
| **3** | Alsace, 2020-21,^6^ | R |  | 2020-2021 | Cross sectional | Regional | Alsace | 11-12 |
| **4** | Apété, Perspect Public Health 2012,^7^ Guinhouya, BEH 2010,^8^ | A A |  | 2005-2008 | Cross sectional | Local | Villeneuve d'Ascq Council | 8-11 |
| **5** | Apouey, Econ Hum Biol 2016,^9^ | A | Enquête Santé soins médicaux (ESSM) | 2002-2003 | Cohort | National | Hexagonal France | 2-17 |
| **6** | Assier de Pompignan, RESP 2006,^10^ | A |  | 2003-2003 | Cohort | DROM | Martinique Island | 5-6 |
| **7** | Auvergne, 2006,^11^ | R |  | 2004-2005 | Cross sectional | Regional | Auvergne | 5-6 |
| **8** | Auvergne, 2009,^12^ | R |  | 2007-2008 | Cross sectional | Regional | Auvergne | 8-9 |
| **9** | Baranne, BEH 2021,^13^ | A |  | 2018-2021 | Cross sectional | Departmental | Val de Marne | 4 |
| **10** | Beau, Arch Ped 2017,^14^ | A | Efemeris | 2006-2013 | Cohort | Departmental | Haute-Garonne | 2 |
| **11** | Bois, Arch Ped 2010,^15^ | A |  | 2004-2005 | Cross sectional | Departmental | Hauts-de-Seine | 3-4 |
| **12** | Bois, RESP 2013,^16^ Bois, BEH 2014,^17^ | A |  | 2010 | Cross sectional | Departmental | Hauts-de-Seine | 3-4 |
| **13** | Bonsergent, BMC Public Health 2012,^18^ Bonsergent, Am J Prev Med 2013,^19^ Bonsergent, Global Health Promot 2013,^20^ Briançon, Trials 2010,^21^ Manneville, Prev Med 2021,^22^ | A A A A A | PRALIMAP | 2006-2007 | Trial | Regional | Lorraine | 14-17 |
| **14** | Bretagne, 2017,^23^ | R | Santé des Jeunes scolarisés en Bretagne | 2017 | Cross sectional | Regional | Bretagne | 11-18 |
| **15** | Caïus, RESP 2002,^24^ | A |  | 2001 | Cross sectional | DROM | Martinique Island | 12-18 |
| **16** | Castetbon, 2004,^25^ Péneau, Int J Ped Obes 2011,^26^ Rolland-Cachera, Int J Obes 2002,^27^ | R A A | CE1-CE2 | 2000 | Cross sectional | National | Aix-Marseille, Caen, Lyon, Nancy-Metz, Nantes, Nice, Orléans-Tours, Paris, Rouen, Toulouse, Versailles | 7-9 |
| **17** | Chardon, 2013,^28^ | R | DRESS - CM2 | 2007-2008 | Cross sectional | National | Hexagonal France + DROM | 10-11 |
| **18** | Chardon, 2014,^29^ | R | DRESS - 3e | 2008-2009 | Cross sectional | National | Hexagonal France + DROM | 14-15 |
| **19** | Chardon, 2015,^30^ | R | DRESS - GS | 2012-2013 | Cross sectional | National | Hexagonal France + DROM (excluding Corse, Bretagne, Mayotte) | 5-6 |
| **20** | Chollet, Arch ped 2013,^31^ | A |  | 2007-2009 | Cross sectional | Departmental | Haute-Garonne | 3-4 |
| **21** | Correze, 2017-19,^32^ | R | Etat de santé de la petite enfance en Corrèze | 2017-2019 | Cross sectional | Departmental | Corrèze | 4-6 |
| **22** | Correze, 2019-20,^33^ | R | Etat de santé de la petite enfance en Corrèze | 2019-2020 | Cross sectional | Departmental | Corrèze | 4-6 |
| **23** | Correze, 2020-21,^34^ | R | Etat de santé de la petite enfance en Corrèze | 2020-2021 | Cross sectional | Departmental | Corrèze | 4-6 |
| **24** | Creuse, 2016-19,^35^ | R | Etat de santé de la petite enfance en Creuse | 2004-2019 | Cross sectional | Departmental | Creuse | 3-6 |
| **25** | Currie, 2004,^36^ Currie, J Ado Health 2012,^37^ Due, Int J Obes 2009,^38^ | R A A | HBSC | 2001-2002 | Cross sectional | National | Hexagonal France | 11-15 |
| **26** | Currie, 2008,^39^ Dupuy, BMC Public Health 2011,^40^  Gaudineau, BMC Pub Health 2010,^41^  Haug, Int J Pub Health 2009,^42^ | R A A A | HBSC | 2005-2006 | Cross sectional | National | Hexagonal France | 11-15 |
| **27** | Currie, 2012,^43^ HBSC, 2009-10,^44^ | R R | HBSC | 2009-2010 | Cross sectional | National | Hexagonal France | 11-15 |
| **28** | Daigre, Diabet Metabl 2012,^45^ | A | PODIUM | 2007-2008 | Cross sectional | DROM-COM | Guadeloupe Island,  Martinique Island,  French Guiana,  French Polynesia |  |
| **29** | De Peretti, 2004,^46^ | R | DRESS - 3e | 2000-2001 | Cross sectional | National | Hexagonal France + DROM | 14-15 |
| **30** | Deheeger, Arch Ped 2004,^47^ | A |  | 2000 | Cohort | Local | Paris | 2-16 |
| **31** | Devaux, Eur J Public Health 2013,^48^ | A | Enquête santé et protection sociale (ESPS) | 2000-2014 | Cross sectional | National | Hexagonal France | 2-17 |
| **32** | Diouf, Epidemiology 2010,^49^ | A | Obepi | 2000-2006 | Cross sectional | National | Hexagonal France | 15-18 |
| **33** | Fernandez, BEH 2007,^50^ Alsace, 6 ans 2001-02,^51^ | A R |  | 2001-2002 | Cross sectional | Regional | Alsace | 5-6 |
| **34** | Feur, BEH 2007a,^52^  Feur, BEH 2007b,^53^ | A A |  | 2005-2005 | Cross sectional | Departmental | Val de Marne | 10-17 |
| **35** | Frayon, BMC Pub Health 2017,^54^ Frayon, Ethnicity Health 2019,^55^ Frayon, Pediatr Obes 2018,^56^ Wattelez, Nutrients 2019,^57^ | A A A A |  | 2015-2016 | Quasi  experimental | COM | New Caledonia | 11-16 |
| **36** | Frayon, Lancet Reg Health 2020,^58^ Galy, Nutrients 2020,^59^ | A A |  | 2018-2019 | Cross sectional | COM | New Caledonia | 10-16 |
| **37** | Gatti, Journal of Adolescent Health 2015,^60^ | A |  | 2011 | Trial | COM | French Polynesia | 10-18 |
| **38** | Ginioux, SFSP 2006,^61^ | A |  | 2003-2004 | Cross sectional | Departmental | Seine Saint Denis | 5-18 |
| **39** | Guignon, 2002,^62^ Guignon, 2003,^63^ Duport, BEH 2003,^64^ Guignon, RESP 2004,^65^ | R R A A | DRESS - GS | 2000-2001 | Cross sectional | National | Hexagonal France + DROM | 5-6 |
| **40** | Guignon, 2007,^66^ Deschamps, Ped Obesity 2015,^67^ | R A | DRESS - 3e | 2003-2004 | Cross sectional | National | Hexagonal France | 14-15 |
| **41** | Guignon, 2008,^68^ | R | DRESS - CM2 | 2004-2005 | Cross sectional | National | Hexagonal France + DROM (excluding French Guiana) | 10-11 |
| **42** | Guignon, 2010,^69^ | R | DRESS - GS | 2005-2006 | Cross sectional | National | Hexagonal France + DROM | 5-6 |
| **43** | Guignon, 2017,^70^ | R | DRESS - CM2 | 2014-2015 | Cross sectional | National | Hexagonal France + DROM | 10-11 |
| **44** | Guignon, 2019,^71^ | R | DRESS - 3e | 2016-2017 | Cross sectional | National | Hexagonal France + DROM | 14-15 |
| **45** | Haute-normandie, 2012-13,^72^ | R | INDIcateurs de Suivi en Santé (Indiss) | 2012-2013 | Cross sectional | Regional | Haute-Normandie | 14-17 |
| **46** | Haute-normandie,2012-13,^73^ | R | INDIcateurs de Suivi en Santé (Indiss) | 2012-2013 | Cross sectional | Regional | Haute-Normandie | 11-14 |
| **47** | Haute-savoie, 200,^74^ | R |  | 2003 | Cross sectional | Departmental | Haute-Savoie | 10-14 |
| **48** | Haute-savoie, 2008,^75^ Fontaine, BEH 2010,^76^ | R A | La santé des enfants de 0-6 ans en Rhône-Alpes | 2007-2008 | Cross sectional | Departmental | Haute-Savoie | 5-6 |
| **49** | Hauts de France, 2018-19,^77^ | R | Jeunes En SAnté Indicateurs et Suivi (Jesais) | 2018-2019 | Cross sectional | Regional | Picardie | 11 |
| **50** | HBSC, 2017-18,^78^ Inchley, 2020,^79^ | R R | HBSC-EnCLASS | 2017-2018 | Cross sectional | National | Hexagonal France | 11-15 |
| **51** | Helfenstein, Obes 2006,^80^ Lorraine, 2001,^81^ | A R |  | 2000 | Cross sectional | Regional | Lorraine | 3-15 |
| **52** | Heude, Diabetes Metab 2003,^82^ | A | FLVS I et II | 2000 | Cross sectional | Local | Fleurbaix and Laventie | 5-11 |
| **53** | Ile de France, 2002-03,^83^ | R | Extension régionale de l’enquete decenale de l’Insee | 2002-2003 | Cross sectional | Regional | Ile de France | 2-18 |
| **54** | INCA, 2009,^84^ Lioret, Br J Nutr 2011,^85^ Lioret, British J Nutr 2010,^86^ Lioret, Obesity 2009,^87^ | R A A A | INCA 2 | 2006-2007 | Cross sectional | National | Hexagonal France (excluding Corse) | 3-17 |
| **55** | INCA, 2017,^87^ | R | INCA 3 | 2014-2015 | Cross sectional | National | Hexagonal France (excluding Corse) | 2-17 |
| **56** | Inchley, 2016,^88^ Ngantcha, J Phys Act Health 2018,^89^ HBSC, 2013-14,^90^ | R A R | HBSC | 2013-2014 | Cross sectional | National | Hexagonal France | 11-15 |
| **57** | SPF, 2007,^91^ | R | ENNS | 2006-2007 | Cross sectional | National | Hexagonal France | 3-17 |
| **58** | SPF, 2017,^92^ | R | ESTEBAN | 2014-2016 | Cross sectional | National | Hexagonal France (excluding Corse) | 6-17 |
| **59** | Jouret, Am J Clin Nutr 2007,^93^ | A | RéPOP | 2002-2003 | Cross sectional | Departmental | Haute-Garonne | 3 |
| **60** | Kêkê, RESP 2015,^94^ | A |  | 2009 | Cross sectional | Local | Maubeuge-Val-de-Sambre | 5-12 |
| **61** | Klein-Platat, Diab Metab Res Rev 2003,^95^ Wagner, Diabetes Metab 2004,^96^ Klein-Platat, Int J Obes 2005,^97^ Casey, Int J Obes 2012,^98^ | A A A A |  | 2001 | Cross sectional | Departmental | Bas-Rhin | 12 |
| **62** | La Réunion, 2006-07,^99^ | R | ETADAR | 2006-2007 | Cross sectional | DROM | La Réunion Island | 13-17 |
| **63** | La Réunion, 2011-12,^100^ | R |  | 2011-2012 | Cross sectional | DROM | La Réunion Island | 4-16 |
| **64** | Labeyrie, 2004,^101^ | R | DRESS - CM2 | 2001-2002 | Cross sectional | National | Hexagonal France + DROM (excluding Clermont-Ferrand, Poitiers) | 10-11 |
| **65** | Lanckriet, Journal of Physical Activity Research 2017,^102^ | A |  | 2015 | Trial | Local | Flandre Lys Towns (Estaires, Merville, Haverskerque et La Gorgue, Fleurbaix, Lestrem, Laventie et Sailly sur la Lys) | 8-11 |
| **66** | Lanfranchi, Scand J Med Sci Sports 2014,^103^ | A |  | 2007-2008 | Cross sectional | Local | Côte d'Azur | 11-18 |
| **67** | Legleye, PloS One 2014,^104^ | A |  | 2010 | Cross sectional | Local | Paris | 17-18 |
| **68** | Limousin, 2013-14,^105^ | R | Etat de santé de la petite enfance dans le Limousin | 2013-2014 | Cross sectional | Regional | Limousin | 9-15 |
| **69** | Limousin, 2015-16,^106^ | R | Etat de santé de la petite enfance dans le Limousin | 2015-2016 | Cross sectional | Regional | Limousin | 10-14 |
| **70** | Limousin, 2016-17,^107^ | R | Etat de santé de la petite enfance dans le Limousin | 2016-2017 | Cross sectional | Regional | Limousin | 10-14 |
| **71** | Lorraine, 2014-15,^108^ | R |  | 2014-2015 | Cross sectional | Regional | Lorraine | 6 |
| **72** | Mayotte, 2006,^109^ | R | NutriMay | 2006 | Cross sectional | DROM | Mayotte | 2-13 |
| **73** | Messaadi, RESP 2020,^110^ | A | EXDEMPAGE | 2015 | Cross sectional | Local | Lille | 11 |
| **74** | Midi-Pyrénées, 2018,^111^ | R | Infiscol | 2017-2018 | Cross sectional | Regional | Occitanie | 5-12 |
| **75** | Milcent, Front Pediatr 2022,^112^  Le Gal, Sci Rep 2023,^113^ | A | ELFE | 2014-2016 | Cohort  Cross sectional | National | Hexagonal France | 3-4 |
| **76** | Moschonis, Eur J Ped 2017,^114^ De Lauzon Guillain, Pediatric Obes 2019,^115^ Maître, BMJ Open 2018,^116^ Ruiz, Paed Perinat Epidemiol 2016,^117^ | A A A A | EDEN | 2009-2012 | Cohort | Local | Nancy  Poitiers | 5-6 |
| **77** | Nantes, 2018,^118^ | R |  | 2012-2022 | Cross sectional | Local | Nantes | 5-9 |
| **78** | Normandie, 2017-18,^119^ | R | Enquête auprès des JEUnes sur la Santé (EnJEU Santé) | 2017-2018 | Cross sectional | Departmental | Eure Seine Maritime | 11 |
| **79** | Normandie, 2018-20,^120^ | R | Enquête auprès des JEUnes sur la Santé (EnJEU Santé) | 2018-2020 | Cross sectional | Regional | Normandie | 11 |
| **80** | Nouvelle-Aquitaine, 2017-18,^121^ | R |  | 2017-2018 | Cross sectional | Regional | Nouvelle Aquitaine (hors Charentes-Maritimes) | 11-12 |
| **81** | Nouvelle-Aquitaine, 2018-21,^122^ | R |  | 2018-2021 | Cross sectional | Regional | Nouvelle Aquitaine | 11-12 |
| **82** | Ottova, Qual Life Res 2012,^123^ | A | KIDSCREEN | 2003 | Cross sectional | National | Hexagonal France | 8-18 |
| **83** | Provence Alpes Cotes d’Azur, 2009-10,^124^ | R | Eval Mater | 2009-2010 | Cross sectional | Regional | Provence-Alpes-Côte d'Azur | 3.5-4.5 |
| **84** | Padilla, Santé Publique 2012,^125^ | A | EPODE | 2008-2009 | Cross sectional | Local | Narbonne | 5-10 |
| **85** | Paineau, Arch Pediatr Adolesc Med 2008,^126^ | A | ELPAS | 2005 | Trial | Local | Paris | 7-9 |
| **86** | Paris, 2009,^127^ | R |  | 2008-2009 | Cross sectional | Local | Paris | 5-9 |
| **87** | Péneau, Int J Obes 2009,^128^ | A | IRSA | 2000-2006 | Cross sectional | Regional | Centre  Pays de la Loire Normandie | 6-15 |
| **88** | Picardie, 2007-08,^129^ | R | Jeunes En SAnté Indicateurs et Suivi (Jesais) | 2007-2008 | Cross sectional | Regional | Picardie | 15 |
| **89** | Picardie, 2009-15,^130^ Picardie, 2009-13,^131^ | R R | Jeunes En SAnté Indicateurs et Suivi (Jesais) | 2008-2015 | Cross sectional | Regional | Picardie | 15 |
| **90** | Picardie, 2015-19,^132^ | R | Jeunes En SAnté Indicateurs et Suivi (Jesais) | 2015-2019 | Cross sectional | Regional | Picardie | 15 |
| **91** | Picardie, 2005-06,^133^ | R | Jeunes En SAnté Indicateurs et Suivi (Jesais) | 2005-2006 | Cross sectional | Regional | Picardie | 11 |
| **92** | Picardie, 2008-15,^134^ Picardie, 2008-13,^135^ | R R | Jeunes En SAnté Indicateurs et Suivi (Jesais) | 2008-2015 | Cross sectional | Regional | Picardie | 11 |
| **93** | Picardie, 2006-07,^136^ | R | Jeunes En SAnté Indicateurs et Suivi (Jesais) | 2006-2007 | Cross sectional | Regional | Picardie | 6 |
| **94** | Pitard, BEH 2003,^137^ | A |  | 2003 | Cross sectional | Regional | Haute-Normandie | 11-15 |
| **95** | Pitrou, Obesity 2010,^138^ | A |  | 2004-2005 | Cross sectional | Regional | Provence-Alpes-Côte d'Azur | 6-11 |
| **96** | Puy de dôme, 2012,^139^ | R |  | 2012 | Cross sectional | Departmental | Puy de Dôme | 3-4 |
| **97** | Rambhojan, Nutr Metab 2015,^140^ | A |  | 2013 | Cross sectional | DROM | Guadeloupe Island | 11-15 |
| **98** | Rhône Alpes, 2020,^141^ | R | La santé des enfants de 0-6 ans en Rhône-Alpes | 2017-2018 | Cross sectional | Regional | Auvergne-Rhône-Alpes | 3-4 |
| **99** | Roche, Br J Nutr 2020,^142^ | A |  | 2008-2011 | Cohort | Departmental | Haute-Saone | 3-7 |
| **100** | Romon, Public Health Nutr 2009,^143^ Heude, Obes Res 2004,^144^ | A A | FLVS III | 2002-2004 | Trial | Local | Fleurbaix  Laventie | 5-11 |
| **101** | Roth, Arch Pediatr 2022,^145^ | A |  | 2017-2020 | Cross sectional | Departmental | Bouches-du-Rhônes | 3,5-4,5 |
| **102** | Salanave, 2011,^146^  Salanave, Int J Ped Obes 2009,^147^ Peneau, Int J Ped Obes (Lond) 2011,^148^ | R  A A | CE1-CE2 | 2007 | Cross sectional | National | Hexagonal France | 7-9 |
| **103** | Salanave, 2018,^149^ | R | CE1-CE2 | 2016 | Cross sectional | National | Hexagonal France | 7-9 |
| **104** | Simon, Diabetes Metab 2006,^150^ Simon, Int J Obes 2004,^151^ Simon, Int J Obes 2008,^152^ Simon, Int J Obes 2014,^153^ Delmas, Obesity 2007,^154^ | A A A A A | ICAPS | 2002 | Cluster randomised controlled trial | Departmental | Bas-Rhin | 11-13 |
| **105** | SPF, 2000,^155^ | R | Barometre Santé | 2000 | Cross sectional | National | Hexagonal France | 12-17 |
| **106** | SPF, 2005,^156^ | R | Barometre Santé | 2005 | Cross sectional | National | Hexagonal France | 12-17 |
| **107** | SPF, 2010,^157^ | R | Barometre Santé | 2010 | Cross sectional | National | Hexagonal France | 15-17 |
| **108** | SPF, 2014,^158^ | R | Barometre Santé | 2014 | Cross sectional | National | Hexagonal France | 15-17 |
| **109** | SPF, 2016,^159^ | R | Barometre Santé | 2016 | Cross sectional | National | Hexagonal France | 15-17 |
| **110** | Stoupa, J Pediatr Endocrinol Metab 2015,^160^ | A |  | 2008-2009 | Cohort | Departmental | Paris | 5-11 |
| **111** | Thibault, Acta paed 2013,^161^ | A |  | 2004-2011 | Cross sectional | Local | Bordeaux | 5-6 |
| **112** | Thibault, Arch Pediatr 2010,^162^ | A |  | 2001-2005 | Retrospective  cohort | Regional | Aquitaine | 5-9 |
| **113** | Thibault, Nutrition 2010,^163^ Carriere, J Physiol Biochem 2013,^164^ | A A | PNNS | 2004-2005 | Cross sectional | Regional | Aquitaine | 7-18 |
| **114** | Thibault, Pub Health Nut 2013,^165^ Carriere, Pub Health Nut 2015,^166^ | A A | PNNS | 2007-2009 | Cross sectional | Regional | Aquitaine | 5-11 |
| **115** | Tramini, Caries Res 2009,^167^ | A |  | 2007 | Cross sectional | Local | Montpellier | 12 |
| **116** | Vanhelst, Clinical nutrition 2017,^168^ Cadenas-Sanchez, J Sci Med Sport 2017,^169^ Flieh, Nutrients 2020,^170^ Martinez-Gomez, Am J Prev Med 2010,^171^ Sichert-Hellert, Pub Health Nutr 2011,^172^ Flieh, Nutrients 2021,^173^ | A A A A A A | HELENA | 2006-2007 | Cohort | Local | Lille | 12,5-17,5 |
| **117** | Vanhelst, J Public Health Nutr 2017,^174^ Vanhelst, Clin Physiol Funct Imaging 2016,^175^ | A A | BOUGE ta santé | 2009-2013 | Trial | National | 16 regions of France | 9-16 |
| **118** | Vanhelst, Ped Obes 2021,^176^ Constant, BMC Public Health 2020,^177^ | A A | Vivons en forme | 2008-2015 | Quasi experimental | Local | Beauvais Meyzieu Royan Douchy-les-Mines Saint-Quentin Vitré | 5-10 |
| **119** | Vanhelst, Pub Health Nutr 2020,^178^  Schipman, BEH 2015,^179^ Barbry, Front Sports Act Living 2022,^180^  Vanhelst, BMC Res Notes 2022,^181^ | A A A A | Diagnoform | 2013-2017 | Cross sectional | National | 14 regions of France | 4-12 |
| **120** | Vanhelst, RESP 2020,^182^ | A | Diagnoform | 2010 | Cohort | Local | Henin-Beaumont Carvin | 5-10 |
| **121** | Verger, RESP 2007,^183^ | A | Eval Mater | 2002-2003 | Cross sectional | Regional | Provence-Alpes-Côte d'Azur | 3.5-4.5 |
| **122** | Viguié, Santé Publique 2002,^184^ | A |  | 2000-2001 | Retrospective  cohort | Local | Grenoble | 4-11 |

A: article; R: report; COM. French overseas collectivity; DROM. overseas departments and regions.

#### Supplemental Table 6 Risk of bias of 122 studies included (red circle: high risk, grey circle: unclear, green risk: low risk).

|  | **Author, publication year** | **Representative  sample of targeted  population** | **Appropriate  recruitment of  participants** | **Appropriate exclusion criteria** | **Low refusal rate or attrition rates or weighting** | **Measured weight and height** | **Conclusion** |
| --- | --- | --- | --- | --- | --- | --- | --- |
| **1** | Aisne, 2003-04,^2^ | ⚫ | ⚫ | ⚫ | ⚫ | ⚫ | ⚫ |
| **2** | Alsace, 2014-15,^3^ Alsace, 2014-16,^4^  Alsace, 2014-17,^5^ | ⚫ | ⚫ | ⚫ | ⚫ | ⚫ | ⚫ |
| **3** | Alsace, 2020-21,^6^ | ⚫ | ⚫ | ⚫ | ⚫ | ⚫ | ⚫ |
| **4** | Apété, Perspect Public Health 2012,^7^ Guinhouya, BEH 2010,^8^ | ⚫ | ⚫ | ⚫ | ⚫ | ⚫ | ⚫ |
| **5** | Apouey, Econ Hum Biol 2016,^9^ | ⚫ | ⚫ | ⚫ | ⚫ | ⚫ | ⚫ |
| **6** | Assier de Pompignan, RESP 2006,^10^ | ⚫ | ⚫ | ⚫ | ⚫ | ⚫ | ⚫ |
| **7** | Auvergne, 2006,^11^ | ⚫ | ⚫ | ⚫ | ⚫ | ⚫ | ⚫ |
| **8** | Auvergne, 2009,^12^ | ⚫ | ⚫ | ⚫ | ⚫ | ⚫ | ⚫ |
| **9** | Baranne, BEH 2021,^13^ | ⚫ | ⚫ | ⚫ | ⚫ | ⚫ | ⚫ |
| **10** | Beau, Arch Ped 2017,^14^ | ⚫ | ⚫ | ⚫ | ⚫ | ⚫ | ⚫ |
| **11** | Bois, Arch Ped 2010,^15^ | ⚫ | ⚫ | ⚫ | ⚫ | ⚫ | ⚫ |
| **12** | Bois, RESP 2013,^16^ Bois, BEH 2014,^17^ | ⚫ | ⚫ | ⚫ | ⚫ | ⚫ | ⚫ |
| **13** | Bonsergent, BMC Public Health 2012,^18^ Bonsergent, Am J Prev Med 2013,^19^ Bonsergent, Global Health Promot 2013,^20^ Briançon, Trials 2010,^21^ Manneville, Prev Med 2021,^22^ | ⚫ | ⚫ | ⚫ | ⚫ | ⚫ | ⚫ |
| **14** | Bretagne, 2017,^23^ | ⚫ | ⚫ | ⚫ | ⚫ | ⚫ | ⚫ |
| **15** | Caïus, RESP 2002,^24^ | ⚫ | ⚫ | ⚫ | ⚫ | ⚫ | ⚫ |
| **16** | Castetbon, 2004,^25^ Péneau, Int J Ped Obes 2011,^26^ Rolland-Cachera, Int J Obes 2002,^27^ | ⚫ | ⚫ | ⚫ | ⚫ | ⚫ | ⚫ |
| **17** | Chardon, 2013,^28^ | ⚫ | ⚫ | ⚫ | ⚫ | ⚫ | ⚫ |
| **18** | Chardon, 2014,^29^ | ⚫ | ⚫ | ⚫ | ⚫ | ⚫ | ⚫ |
| **19** | Chardon, 2015,^30^ | ⚫ | ⚫ | ⚫ | ⚫ | ⚫ | ⚫ |
| **20** | Chollet, Arch ped 2013,^31^ | ⚫ | ⚫ | ⚫ | ⚫ | ⚫ | ⚫ |
| **21** | Correze, 2017-19,^32^ | ⚫ | ⚫ | ⚫ | ⚫ | ⚫ | ⚫ |
| **22** | Correze, 2019-20,^33^ | ⚫ | ⚫ | ⚫ | ⚫ | ⚫ | ⚫ |
| **23** | Correze, 2020-21,^34^ | ⚫ | ⚫ | ⚫ | ⚫ | ⚫ | ⚫ |
| **24** | Creuse, 2016-19,^35^ | ⚫ | ⚫ | ⚫ | ⚫ | ⚫ | ⚫ |
| **25** | Currie, 2004,^36^ Currie, J Ado Health 2012,^37^ Due, Int J Obes 2009,^38^ | ⚫ | ⚫ | ⚫ | ⚫ | ⚫ | ⚫ |
| **26** | Currie, 2008,^39^ Dupuy, BMC Public Health 2011,^40^  Gaudineau, BMC Pub Health 2010,^41^  Haug, Int J Pub Health 2009,^42^ | ⚫ | ⚫ | ⚫ | ⚫ | ⚫ | ⚫ |
| **27** | Currie, 2012,^43^ HBSC, 2009-10,^44^ | ⚫ | ⚫ | ⚫ | ⚫ | ⚫ | ⚫ |
| **28** | Daigre, Diabet Metabl 2012,^45^ | ⚫ | ⚫ | ⚫ | ⚫ | ⚫ | ⚫ |
| **29** | De Peretti, 2004,^46^ | ⚫ | ⚫ | ⚫ | ⚫ | ⚫ | ⚫ |
| **30** | Deheeger, Arch Ped 2004,^47^ | ⚫ | ⚫ | ⚫ | ⚫ | ⚫ | ⚫ |
| **31** | Devaux, Eur J Public Health 2013,^48^ | ⚫ | ⚫ | ⚫ | ⚫ | ⚫ | ⚫ |
| **32** | Diouf, Epidemiology 2010,^49^ | ⚫ | ⚫ | ⚫ | ⚫ | ⚫ | ⚫ |
| **33** | Fernandez, BEH 2007,^50^ Alsace, 6 ans 2001-02,^51^ | ⚫ | ⚫ | ⚫ | ⚫ | ⚫ | ⚫ |
| **34** | Feur, BEH 2007a,^52^  Feur, BEH 2007b,^53^ | ⚫ | ⚫ | ⚫ | ⚫ | ⚫ | ⚫ |
| **35** | Frayon, BMC Pub Health 2017,^54^ Frayon, Ethnicity Health 2019,^55^ Frayon, Pediatr Obes 2018,^56^ Wattelez, Nutrients 2019,^57^ | ⚫ | ⚫ | ⚫ | ⚫ | ⚫ | ⚫ |
| **36** | Frayon, Lancet Reg Health 2020,^58^ Galy, Nutrients 2020,^59^ | ⚫ | ⚫ | ⚫ | ⚫ | ⚫ | ⚫ |
| **37** | Gatti, Journal of Adolescent Health 2015,^60^ | ⚫ | ⚫ | ⚫ | ⚫ | ⚫ | ⚫ |
| **38** | Ginioux, SFSP 2006,^61^ | ⚫ | ⚫ | ⚫ | ⚫ | ⚫ | ⚫ |
| **39** | Guignon, 2002,^62^ Guignon, 2003,^63^ Duport, BEH 2003,^64^ Guignon, RESP 2004,^65^ | ⚫ | ⚫ | ⚫ | ⚫ | ⚫ | ⚫ |
| **40** | Guignon, 2007,^66^ Deschamps, Ped Obesity 2015,^67^ | ⚫ | ⚫ | ⚫ | ⚫ | ⚫ | ⚫ |
| **41** | Guignon, 2008,^68^ | ⚫ | ⚫ | ⚫ | ⚫ | ⚫ | ⚫ |
| **42** | Guignon, 2010,^69^ | ⚫ | ⚫ | ⚫ | ⚫ | ⚫ | ⚫ |
| **43** | Guignon, 2017,^70^ | ⚫ | ⚫ | ⚫ | ⚫ | ⚫ | ⚫ |
| **44** | Guignon, 2019,^71^ | ⚫ | ⚫ | ⚫ | ⚫ | ⚫ | ⚫ |
| **45** | Haute-normandie, 2012-13,^72^ | ⚫ | ⚫ | ⚫ | ⚫ | ⚫ | ⚫ |
| **46** | Haute-normandie,2012-13,^73^ | ⚫ | ⚫ | ⚫ | ⚫ | ⚫ | ⚫ |
| **47** | Haute-savoie, 200,^74^ | ⚫ | ⚫ | ⚫ | ⚫ | ⚫ | ⚫ |
| **48** | Haute-savoie, 2008,^75^ Fontaine, BEH 2010,^76^ | ⚫ | ⚫ | ⚫ | ⚫ | ⚫ | ⚫ |
| **49** | Hauts de France, 2018-19,^77^ | ⚫ | ⚫ | ⚫ | ⚫ | ⚫ | ⚫ |
| **50** | HBSC, 2017-18,^78^ Inchley, 2020,^79^ | ⚫ | ⚫ | ⚫ | ⚫ | ⚫ | ⚫ |
| **51** | Helfenstein, Obes 2006,^80^ Lorraine, 2001,^81^ | ⚫ | ⚫ | ⚫ | ⚫ | ⚫ | ⚫ |
| **52** | Heude, Diabetes Metab 2003,^82^ | ⚫ | ⚫ | ⚫ | ⚫ | ⚫ | ⚫ |
| **53** | Ile de France, 2002-03,^83^ | ⚫ | ⚫ | ⚫ | ⚫ | ⚫ | ⚫ |
| **54** | INCA, 2009,^84^ Lioret, Br J Nutr 2011,^85^ Lioret, British J Nutr 2010,^86^ Lioret, Obesity 2009,^87^ | ⚫ | ⚫ | ⚫ | ⚫ | ⚫ | ⚫ |
| **55** | INCA, 2017,^87^ | ⚫ | ⚫ | ⚫ | ⚫ | ⚫ | ⚫ |
| **56** | Inchley, 2016,^88^ Ngantcha, J Phys Act Health 2018,^89^ HBSC, 2013-14,^90^ | ⚫ | ⚫ | ⚫ | ⚫ | ⚫ | ⚫ |
| **57** | SPF, 2007,^91^ | ⚫ | ⚫ | ⚫ | ⚫ | ⚫ | ⚫ |
| **58** | SPF, 2017,^92^ | ⚫ | ⚫ | ⚫ | ⚫ | ⚫ | ⚫ |
| **59** | Jouret, Am J Clin Nutr 2007,^93^ | ⚫ | ⚫ | ⚫ | ⚫ | ⚫ | ⚫ |
| **60** | Kêkê, RESP 2015,^94^ | ⚫ | ⚫ | ⚫ | ⚫ | ⚫ | ⚫ |
| **61** | Klein-Platat, Diab Metab Res Rev 2003,^95^ Wagner, Diabetes Metab 2004,^96^ Klein-Platat, Int J Obes 2005,^97^ Casey, Int J Obes 2012,^98^ | ⚫ | ⚫ | ⚫ | ⚫ | ⚫ | ⚫ |
| **62** | La Réunion, 2006-07,^99^ | ⚫ | ⚫ | ⚫ | ⚫ | ⚫ | ⚫ |
| **63** | La Réunion, 2011-12,^100^ | ⚫ | ⚫ | ⚫ | ⚫ | ⚫ | ⚫ |
| **64** | Labeyrie, 2004,^101^ | ⚫ | ⚫ | ⚫ | ⚫ | ⚫ | ⚫ |
| **65** | Lanckriet, Journal of Physical Activity Research 2017,^102^ | ⚫ | ⚫ | ⚫ | ⚫ | ⚫ | ⚫ |
| **66** | Lanfranchi, Scand J Med Sci Sports 2014,^103^ | ⚫ | ⚫ | ⚫ | ⚫ | ⚫ | ⚫ |
| **67** | Legleye, PloS One 2014,^104^ | ⚫ | ⚫ | ⚫ | ⚫ | ⚫ | ⚫ |
| **68** | Limousin, 2013-14,^105^ | ⚫ | ⚫ | ⚫ | ⚫ | ⚫ | ⚫ |
| **69** | Limousin, 2015-16,^106^ | ⚫ | ⚫ | ⚫ | ⚫ | ⚫ | ⚫ |
| **70** | Limousin, 2016-17,^107^ | ⚫ | ⚫ | ⚫ | ⚫ | ⚫ | ⚫ |
| **71** | Lorraine, 2014-15,^108^ | ⚫ | ⚫ | ⚫ | ⚫ | ⚫ | ⚫ |
| **72** | Mayotte, 2006,^109^ | ⚫ | ⚫ | ⚫ | ⚫ | ⚫ | ⚫ |
| **73** | Messaadi, RESP 2020,^110^ | ⚫ | ⚫ | ⚫ | ⚫ | ⚫ | ⚫ |
| **74** | Midi-Pyrénées, 2018,^111^ | ⚫ | ⚫ | ⚫ | ⚫ | ⚫ | ⚫ |
| **75** | Milcent, Front Pediatr 2022,^112^  Le Gal, Sci Rep 2023,^113^ | ⚫ | ⚫ | ⚫ | ⚫ | ⚫ | ⚫ |
| **76** | Moschonis, Eur J Ped 2017,^114^ De Lauzon Guillain, Pediatric Obes 2019,^115^ Maître, BMJ Open 2018,^116^ Ruiz, Paed Perinat Epidemiol 2016,^117^ | ⚫ | ⚫ | ⚫ | ⚫ | ⚫ | ⚫ |
| **77** | Nantes, 2018,^118^ | ⚫ | ⚫ | ⚫ | ⚫ | ⚫ | ⚫ |
| **78** | Normandie, 2017-18,^119^ | ⚫ | ⚫ | ⚫ | ⚫ | ⚫ | ⚫ |
| **79** | Normandie, 2018-20,^120^ | ⚫ | ⚫ | ⚫ | ⚫ | ⚫ | ⚫ |
| **80** | Nouvelle-Aquitaine, 2017-18,^121^ | ⚫ | ⚫ | ⚫ | ⚫ | ⚫ | ⚫ |
| **81** | Nouvelle-Aquitaine, 2018-21,^122^ | ⚫ | ⚫ | ⚫ | ⚫ | ⚫ | ⚫ |
| **82** | Ottova, Qual Life Res 2012,^123^ | ⚫ | ⚫ | ⚫ | ⚫ | ⚫ | ⚫ |
| **83** | Provence Alpes Cotes d’Azur, 2009-10,^124^ | ⚫ | ⚫ | ⚫ | ⚫ | ⚫ | ⚫ |
| **84** | Padilla, Santé Publique 2012,^125^ | ⚫ | ⚫ | ⚫ | ⚫ | ⚫ | ⚫ |
| **85** | Paineau, Arch Pediatr Adolesc Med 2008,^126^ | ⚫ | ⚫ | ⚫ | ⚫ | ⚫ | ⚫ |
| **86** | Paris, 2009,^127^ | ⚫ | ⚫ | ⚫ | ⚫ | ⚫ | ⚫ |
| **87** | Péneau, Int J Obes 2009,^128^ | ⚫ | ⚫ | ⚫ | ⚫ | ⚫ | ⚫ |
| **88** | Picardie, 2007-08,^129^ | ⚫ | ⚫ | ⚫ | ⚫ | ⚫ | ⚫ |
| **89** | Picardie, 2009-15,^130^ Picardie, 2009-13,^131^ | ⚫ | ⚫ | ⚫ | ⚫ | ⚫ | ⚫ |
| **90** | Picardie, 2015-19,^132^ | ⚫ | ⚫ | ⚫ | ⚫ | ⚫ | ⚫ |
| **91** | Picardie, 2005-06,^133^ | ⚫ | ⚫ | ⚫ | ⚫ | ⚫ | ⚫ |
| **92** | Picardie, 2008-15,^134^ Picardie, 2008-13,^135^ | ⚫ | ⚫ | ⚫ | ⚫ | ⚫ | ⚫ |
| **93** | Picardie, 2006-07,^136^ | ⚫ | ⚫ | ⚫ | ⚫ | ⚫ | ⚫ |
| **94** | Pitard, BEH 2003,^137^ | ⚫ | ⚫ | ⚫ | ⚫ | ⚫ | ⚫ |
| **95** | Pitrou, Obesity 2010,^138^ | ⚫ | ⚫ | ⚫ | ⚫ | ⚫ | ⚫ |
| **96** | Puy de dôme, 2012,^139^ | ⚫ | ⚫ | ⚫ | ⚫ | ⚫ | ⚫ |
| **97** | Rambhojan, Nutr Metab 2015,^140^ | ⚫ | ⚫ | ⚫ | ⚫ | ⚫ | ⚫ |
| **98** | Rhône Alpes, 2020,^141^ | ⚫ | ⚫ | ⚫ | ⚫ | ⚫ | ⚫ |
| **99** | Roche, Br J Nutr 2020,^142^ | ⚫ | ⚫ | ⚫ | ⚫ | ⚫ | ⚫ |
| **100** | Romon, Public Health Nutr 2009,^143^ Heude, Obes Res 2004,^144^ | ⚫ | ⚫ | ⚫ | ⚫ | ⚫ | ⚫ |
| **101** | Roth, Arch Pediatr 2022,^145^ | ⚫ | ⚫ | ⚫ | ⚫ | ⚫ | ⚫ |
| **102** | Salanave, 2011,^146^  Salanave, Int J Ped Obes 2009,^147^ Peneau, Int J Ped Obes (Lond) 2011,^148^ | ⚫ | ⚫ | ⚫ | ⚫ | ⚫ | ⚫ |
| **103** | Salanave, 2018,^149^ | ⚫ | ⚫ | ⚫ | ⚫ | ⚫ | ⚫ |
| **104** | Simon, Diabetes Metab 2006,^150^ Simon, Int J Obes 2004,^151^ Simon, Int J Obes 2008,^152^ Simon, Int J Obes 2014,^153^ Delmas, Obesity 2007,^154^ | ⚫ | ⚫ | ⚫ | ⚫ | ⚫ | ⚫ |
| **105** | SPF, 2000,^155^ | ⚫ | ⚫ | ⚫ | ⚫ | ⚫ | ⚫ |
| **106** | SPF, 2005,^156^ | ⚫ | ⚫ | ⚫ | ⚫ | ⚫ | ⚫ |
| **107** | SPF, 2010,^157^ | ⚫ | ⚫ | ⚫ | ⚫ | ⚫ | ⚫ |
| **108** | SPF, 2014,^158^ | ⚫ | ⚫ | ⚫ | ⚫ | ⚫ | ⚫ |
| **109** | SPF, 2016,^159^ | ⚫ | ⚫ | ⚫ | ⚫ | ⚫ | ⚫ |
| **110** | Stoupa, J Pediatr Endocrinol Metab 2015,^160^ | ⚫ | ⚫ | ⚫ | ⚫ | ⚫ | ⚫ |
| **111** | Thibault, Acta paed 2013,^161^ | ⚫ | ⚫ | ⚫ | ⚫ | ⚫ | ⚫ |
| **112** | Thibault, Arch Pediatr 2010,^162^ | ⚫ | ⚫ | ⚫ | ⚫ | ⚫ | ⚫ |
| **113** | Thibault, Nutrition 2010,^163^ Carriere, J Physiol Biochem 2013,^164^ | ⚫ | ⚫ | ⚫ | ⚫ | ⚫ | ⚫ |
| **114** | Thibault, Pub Health Nut 2013,^165^ Carriere, Pub Health Nut 2015,^166^ | ⚫ | ⚫ | ⚫ | ⚫ | ⚫ | ⚫ |
| **115** | Tramini, Caries Res 2009,^167^ | ⚫ | ⚫ | ⚫ | ⚫ | ⚫ | ⚫ |
| **116** | Vanhelst, Clinical nutrition 2017,^168^ Cadenas-Sanchez, J Sci Med Sport 2017,^169^ Flieh, Nutrients 2020,^170^ Martinez-Gomez, Am J Prev Med 2010,^171^ Sichert-Hellert, Pub Health Nutr 2011,^172^ Flieh, Nutrients 2021,^173^ | ⚫ | ⚫ | ⚫ | ⚫ | ⚫ | ⚫ |
| **117** | Vanhelst, J Public Health Nutr 2017,^174^ Vanhelst, Clin Physiol Funct Imaging 2016,^175^ | ⚫ | ⚫ | ⚫ | ⚫ | ⚫ | ⚫ |
| **118** | Vanhelst, Ped Obes 2021,^176^ Constant, BMC Public Health 2020,^177^ | ⚫ | ⚫ | ⚫ | ⚫ | ⚫ | ⚫ |
| **119** | Vanhelst, Pub Health Nutr 2020,^178^  Schipman, BEH 2015,^179^ Barbry, Front Sports Act Living 2022,^180^  Vanhelst, BMC Res Notes 2022,^181^ | ⚫ | ⚫ | ⚫ | ⚫ | ⚫ | ⚫ |
| **120** | Vanhelst, RESP 2020,^182^ | ⚫ | ⚫ | ⚫ | ⚫ | ⚫ | ⚫ |
| **121** | Verger, RESP 2007,^183^ | ⚫ | ⚫ | ⚫ | ⚫ | ⚫ | ⚫ |
| **122** | Viguié, Santé Publique 2002,^184^ | ⚫ | ⚫ | ⚫ | ⚫ | ⚫ | ⚫ |

#### Supplemental Table 7 Meta-regression results with logit transformation. Temporal trends in the prevalence of overweight (including obesity) and obesity in girls from 2000 to 2017 in France. No study was available after 2017.

|  | **National low-risk studies (n=18, k=28)** | | |  | **All studies^*^** | | |
| --- | --- | --- | --- | --- | --- | --- | --- |
|  | *β* | *[95% CI]* | *P-value* |  | *β* | *[95% CI]* | *P-value* |
| **Overweight (including obesity)** |  |  |  |  |  |  |  |
| Intercept | 14.94 | [12.54; 17.70] |  |  | 12.14 | [11.04; 13.34] |  |
| Calendar year midrange | -0.08 | [-0.24; 0.13] | 0.4027 |  | 0.10 | [0.04; 0.18] | **0.0013** |
| Age class |  |  |  |  |  |  |  |
| *2 to <6* |  | *ref* | **< 0.0001** |  |  | ref | **< 0.0001** |
| *6 to <11* | 7.32 | [3.55; 12.25] |  |  | 3.22 | [2.32; 4.26] |  |
| *11 to <18* | 2.74 | [-0.01; 6.65] |  |  | 3.69 | [2.61; 4.96] |  |
| **Obesity** |  |  |  |  |  |  |  |
| Intercept | 3.80 | [3.08; 4.68] |  |  | 2.61 | [2.22; 3.06] |  |
| Calendar year midrange | 0.02 | [-0.03; 0.10] | 0.5113 |  | 0.06 | [0.03; 0.11] | **< 0.0001** |
| Age class |  |  |  |  |  |  |  |
| *2 to <6* |  | *ref* | 0.1186 |  |  | ref | **0.0087** |
| *6 to <11* | 0.86 | [-0.04; 2.39] |  |  | 0.42 | [0.12; 0.85] |  |
| *11 to <18* | 0.21 | [-0.51; 1.51] |  |  | 0.20 | [-0.10; 0.64] |  |

k: statistical unit. The β estimates and their 95% CIs were back-transformed by using the Freeman-Tukey double arcsine.

^*^ For overweight (including obesity): n=97 studies with data available and k=215; for obesity: n=95 with data available and k=213.

#### Supplemental Table 8 Meta-regression results with logit transformation. Temporal trends in the prevalence of overweight (including obesity) and obesity in boys from 2000 to 2017 in France. No study was available after 2017.

|  | **National low-risk of bias (n=18, k=28)** | | |  | **All studies^*^** | | |
| --- | --- | --- | --- | --- | --- | --- | --- |
|  | *β* | *[95% CI]* | *P-value* |  | *β* | *[95% CI]* | *P-value* |
| **Overweight (including obesity)** |  |  |  |  |  |  |  |
| Intercept | 12.29 | [10.09; 14.89] |  |  | 10.51 | [9.57; 11.53] |  |
| Calendar year midrange | -0.12 | [-0.25; 0.07] | 0.1918 |  | -0.01 | [-0.06; 0.05] | 0.6842 |
| Age class |  |  |  |  |  |  |  |
| *2 to <6* |  | *ref* | **< 0.0001** |  |  |  | **<0 .0001** |
| *6 to <11* | 6.82 | [3.12; 11.94] |  |  | 3.54 | [2.63; 4.60] |  |
| *11 to <18* | 5.70 | [2.27; 10.55] |  |  | 7.26 | [5.80; 8.92] |  |
| **Obesity** |  |  |  |  |  |  |  |
| Intercept | 3.14 | [2.69; 3.66] |  |  | 2.15 | [1.81; 2.54] |  |
| Calendar year midrange | -0.01 | [-0.04; 0.03] | 0.6535 |  | 0.03 | [0.01; 0.06] | **0.0081** |
| Age class |  |  |  |  |  |  |  |
| *2 to <6* |  | *ref* | **< 0.0001** |  |  |  | **< 0.0001** |
| *6 to <11* | 1.33 | [0.55; 2.46] |  |  | 0.61 | [ 0.29; 1.09] |  |
| *11 to <18* | 1.37 | [0.55; 2.59] |  |  | 1.30 | [0.76; 2.06] |  |

k: statistical unit. The β estimates and their 95% CIs were back-transformed by using the Freeman-Tukey double arcsine.

^*^ For overweight (including obesity): n=97 studies with data available and k=215; for obesity: n=95 with data available and k=213.

#### Supplemental Figure 1 Geographic distribution of included studies (n=122).

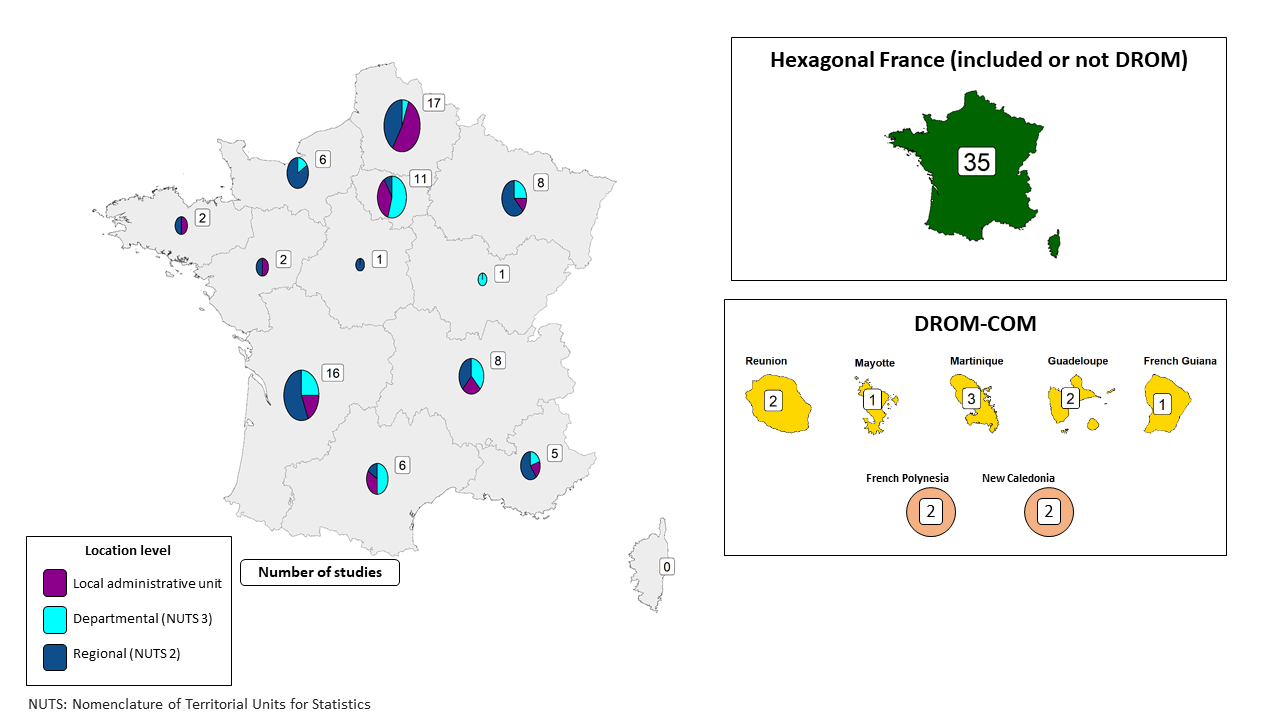

COM. French overseas collectivities; DROM. overseas departments and regions.
Nota Bene. One study can contribute to several French regions.

#### Supplemental Figure 2 Meta-analysis results with logit transformation. Forest plot of 18 nationwide studies at low risk of bias reporting the prevalence of overweight (including obesity) in girls (A) and boys (B), globally and by age class.

A)
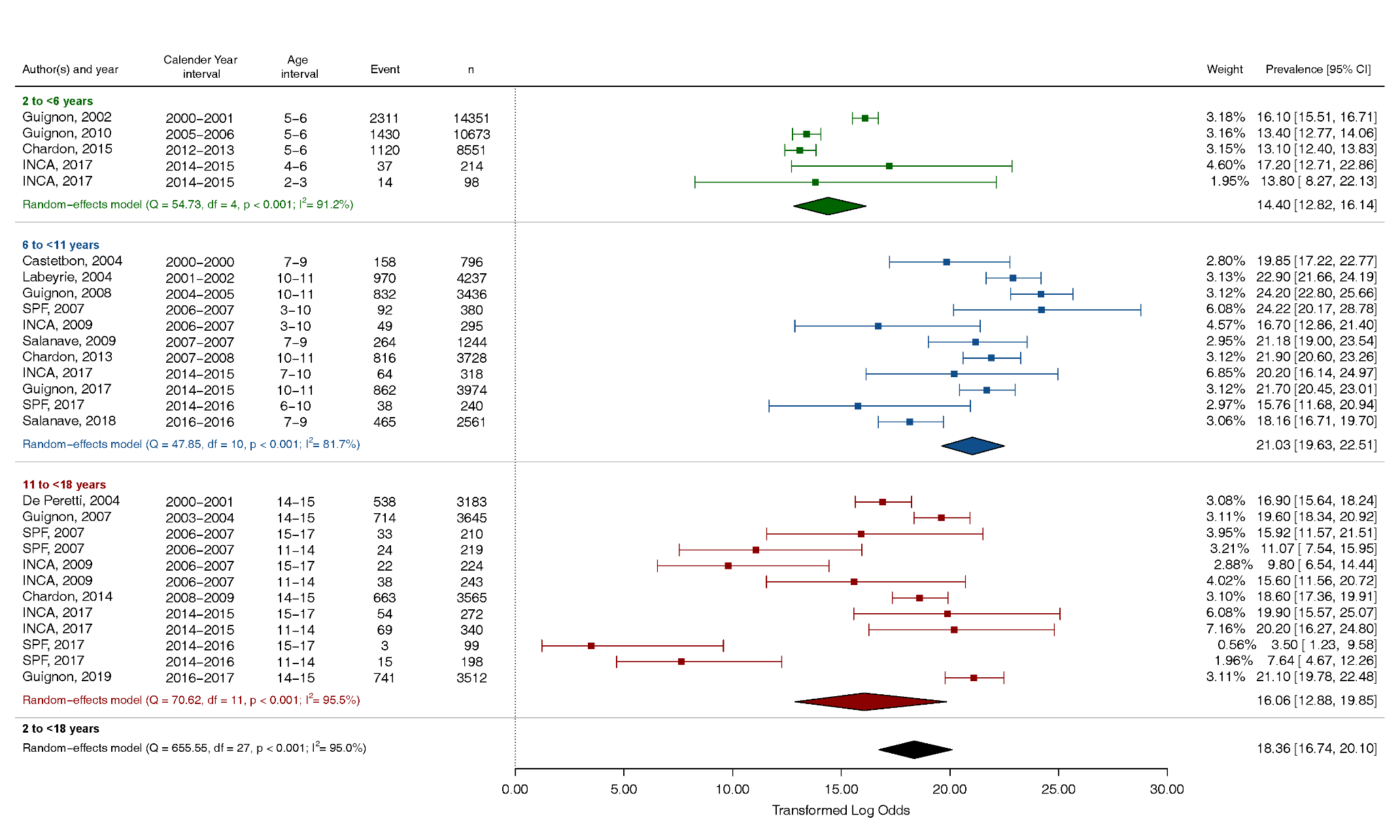

B)
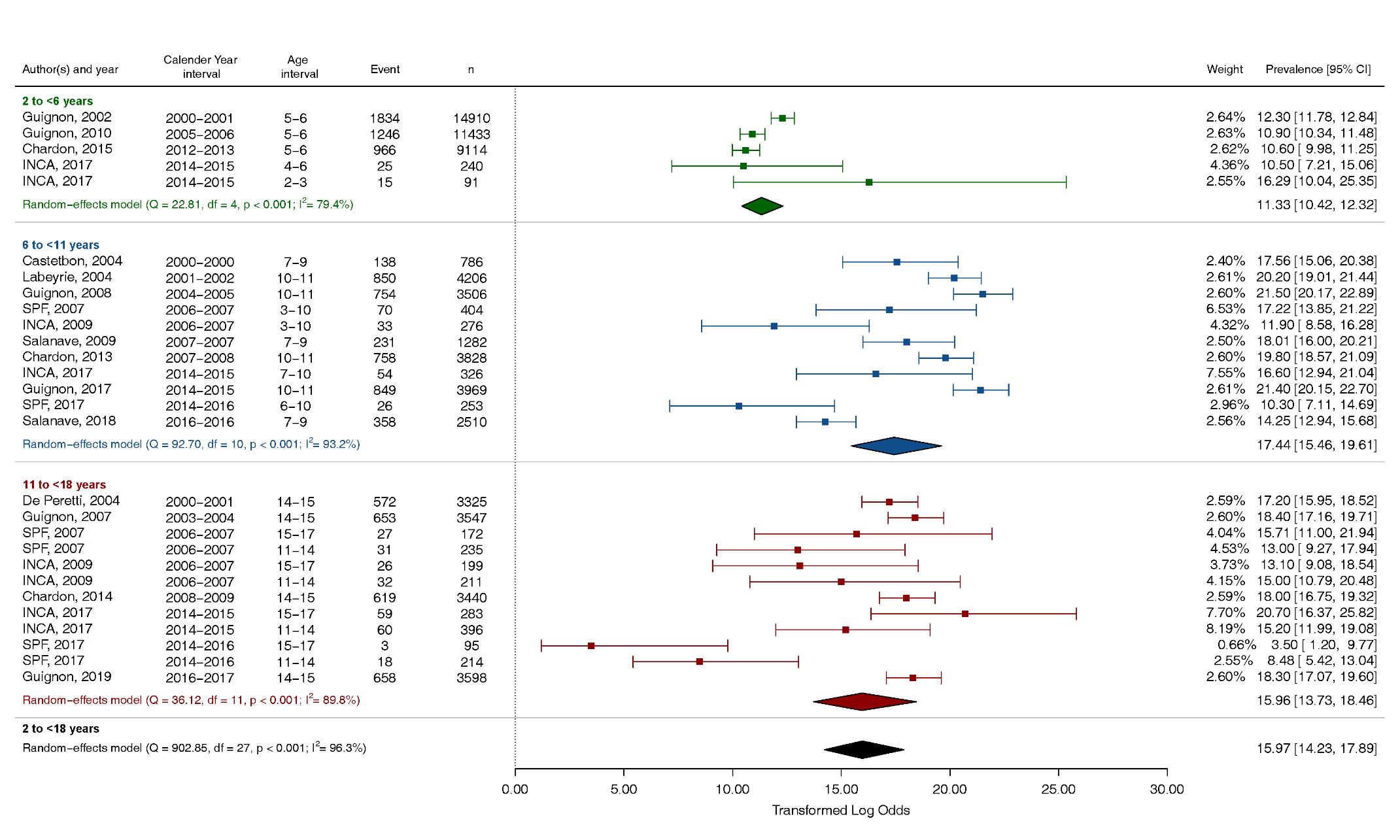

#### Supplemental Figure 3 Meta-analysis results with logit transformation. Forest plot of 18 nationwide studies at low risk of bias reporting the prevalence of obesity in girls (A) and boys (B), globally and by age class.

A)
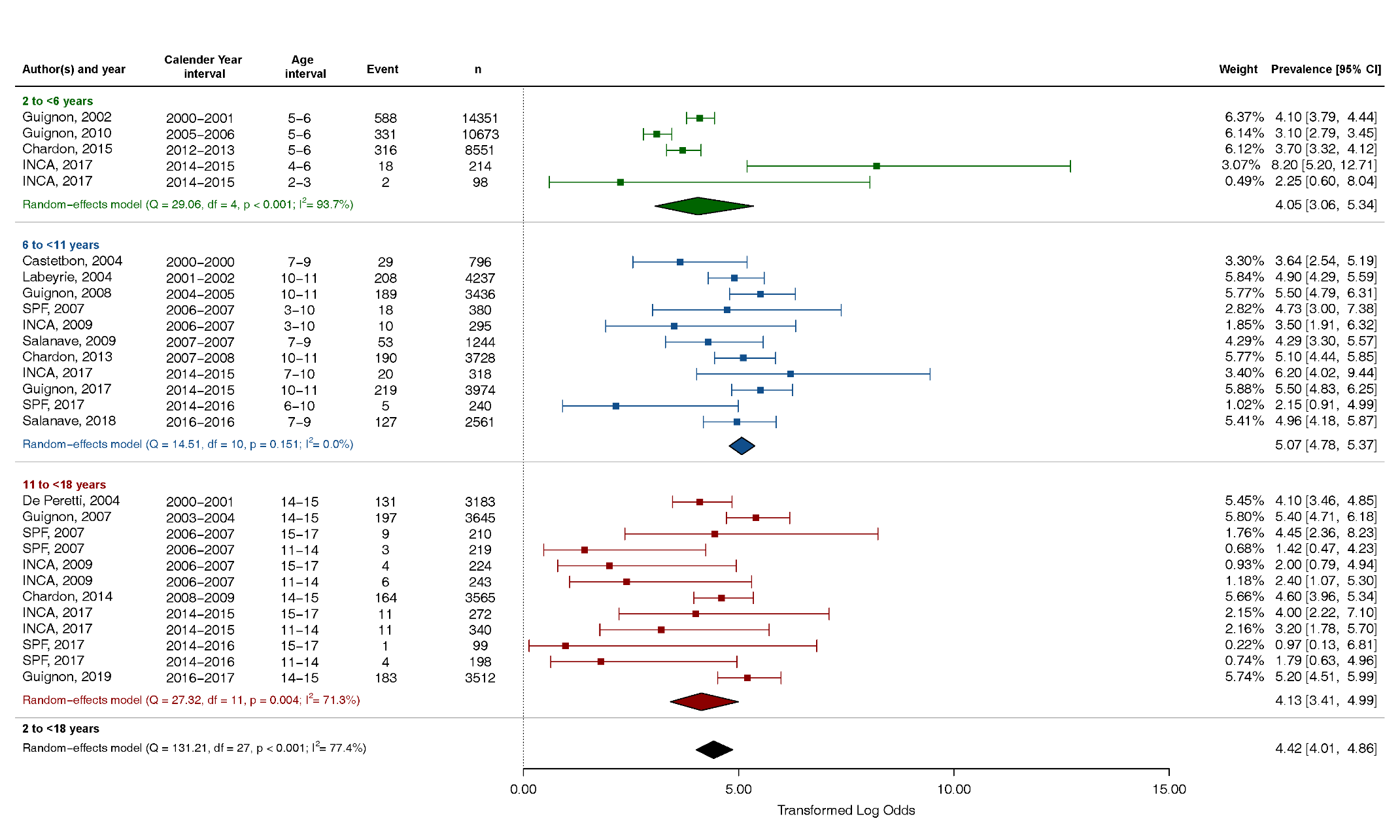

B)
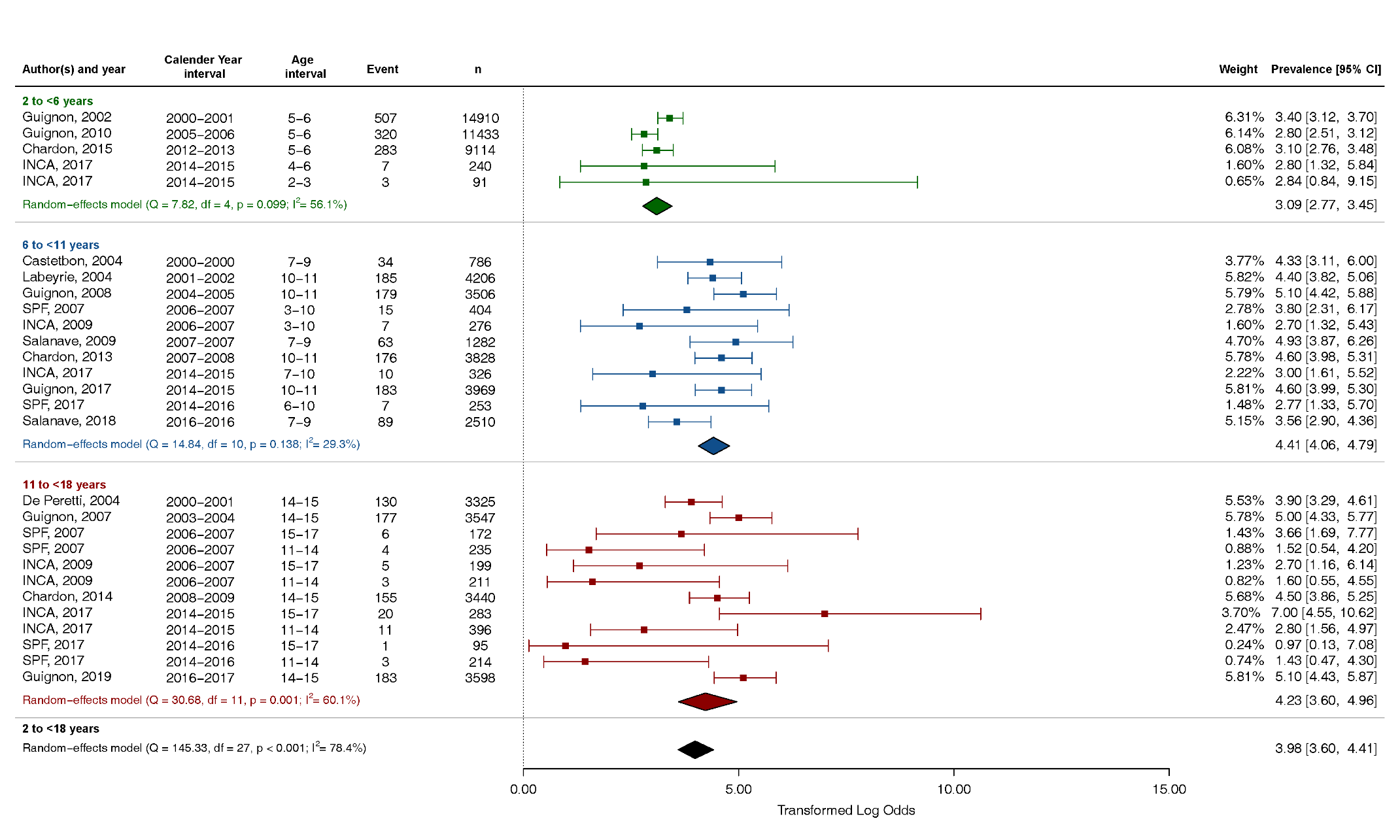

### REFERENCES
